# Epidemiology-Informed Graph Neural Networks for Predicting and Interpreting Transmissible Hospital-Acquired Infections: A Retrospective Cohort and Simulation Study

**DOI:** 10.64898/2026.05.08.26352740

**Authors:** Yamil Vindas, Alban Bornet, Mohamed Abbas, Damien Geissbühler, Jose F. Rodrigues-Jr, Douglas Teodoro

## Abstract

Transmissible hospital-acquired infections (HAIs) arise from complex, time-varying interactions among patients, healthcare workers, and clinical environments. Although data-driven approaches like graph neural networks (GNNs) effectively model these contacts, they often function as black boxes that over-look established epidemiological principles, limiting interpretability and clinical trust. Inspired by physics-informed neural networks, we propose a epidemiology-informed GNN (EIGNN) framework for patient-level state transitions prediction in dynamic hospital settings, integrating mechanistic epidemiological models into GNNs in a principled manner. Patient-level risk factors learned from dynamic contact networks are jointly leveraged to infer latent epidemiological states, predict state transitions across multiple horizons, and estimate key epidemiological parameters, including transmission and recovery rates. We evaluate the approach on a real-world hospital-onset COVID-19 cohort and two public datasets simulating viral and bacterial HAIs. Across multiple architectures and horizons, EIGNNs achieves AUC-ROC up to 98.46% while providing interpretable, mechanistically consistent insights, offering a transparent tool for infection prevention and control.

---

Infection risk assessment plays a central role in infection prevention and control (IPC) strategies. Early identification of high-risk patients enables targeted interventions such as patient isolation, carrier screening, or optimized environmental cleaning, reducing transmissible hospital-acquired infections (HAIs) and supporting stewardship through judicious antibiotic use and more efficient allocation of healthcare resources [1–3]. However, estimating infection risks remains challenging due to the stochastic and non-linear nature of hospital transmission, arising from complex, time-varying interactions between patients, healthcare workers, and clinical environments [4].

Graph neural networks (GNNs) provide a principled framework for learning from graph-structured data and are widely used for modeling dynamic systems [5, 6]. In hospital settings, interactions can be represented as dynamic spatio-temporal heterogeneous graphs, where nodes (patients, healthcare workers) and edges (contacts) evolve over time [7, 8].

Early work focused on discrete-time snapshot-based formulations combining graph convolutions with recurrent or attention-based temporal modeling [9–12]. Subsequent architectures introduced adaptive graph learning and scalability to better capture evolving spatial dependencies [13–16]. To model irregular interactions, continuous-time and event-driven approaches used memory-based mechanisms to capture long-range temporal dependencies [17, 18]. Recent work explored causal and higher-order temporal modeling through De Bruijn Graph Neural Networks and attention-based architectures such as Todyformer and TIDFormer [19–21]. These mechanisms have proven effective in epidemic modeling, enabling focus on high-risk location-specific dynamics [22], with strong performance for predicting colonization status [8]. Further advances in dynamic embeddings and spatio-temporal convolutions highlighted the importance of temporal smoothness and structural dependencies [23–29]. Higher-order extensions show that modeling network motifs and neighborhood overlap improves predictive performance [30]. Despite these advances, purely data-driven methods often lack mechanistic constraints, limiting interpretability and trustworthiness for clinical decision-making.

To address this, epidemiology-informed learning has emerged, building on physics-informed neural networks (PINNs) [31] to embed governing equations and mechanistic constraints into data-driven models. Some approaches incorporate epidemiological ordinary differential equation (ODE) constraints via automatic differentiation [32–36], though these can be sensitive to noise and data sparsity [37, 38]. Conversely, discretize formulations improve interpretability and numerical stability by enforcing explicit state transitions [39–42]. These epidemiology-informed neural networks (EINNs) integrate domain knowledge either as regularization terms [32–34, 36, 40, 43, 44] or directly within model architectures and training objectives [39, 41, 42, 45–50].

Current epidemiology-aware approaches embed mechanistic priors directly into model design, including methods for estimating contact rates [45], hybrid GNN–statistical models with uncertainty quantification [51], next-generation matrix formulations [46], policy-aware spatio-temporal architectures [52], heat-diffusion–based transmission modeling [43], and continuous-time epidemic-aware neural ODEs [50].

Despite this progress, most EINNs target population-level forecasting rather than patient-level infection risk [47–49]. To address this gap, we introduce a network-based, epidemiology-informed graph neural network framework for interpretable patient-level infection risk prediction in dynamic hospital networks. Our approach is architecture-agnostic and embeds mechanistic SIR, SEIR, and SEIRD-NS dynamics within a physics-informed learning paradigm. A GNN first computes node embeddings from dynamic contact networks, which are then used to predict epidemiological states and infer key parameters—such as transmission and recovery rates—by enforcing consistency with governing differential equations via automatic differentiation or discrete-time approximations, under biologically plausible constraints. We evaluate clinical applicability on a real-world COVID-19 hospital cohort and assess mechanistic robustness using two synthetic datasets simulating viral and bacterial HAIs. Our approach matches or outperforms non-informed models while enhancing interpretability and providing insights into pathogen dynamics.

## Results

### Epidemiology-Informed Graph Neural Networks for Patient-Level State Transitions Prediction

Deep neural networks are typically trained by minimizing a task-specific loss. For classification tasks—such as predicting individual state transitions within a forecast window *W* —this is commonly the *cross-entropy* (CE). In PINNs, this objective is augmented with additional terms, often unsupervised, encoding known physical constraints.

Here, we adopt an epidemiology-informed framework in which these constraints are derived from ODEs governing classical models, including SIR with immigration/emigration, SEIR, and SEIRD-NS. The overall training objective is

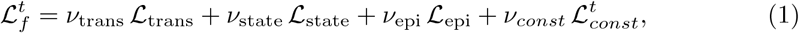

where ℒ_trans_ and ℒ_state_ are supervised losses for transition and state estimation, while ℒ_epi_ and 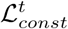 enforce unsupervised consistency with epidemiological ODEs and transition constraints. Formal definitions are provided in Section 5.

Within this framework, GNNs learn per-individual representations from dynamic contact networks to predict patient-level state transitions. The epidemiology-informed formulation further enables joint, unsupervised estimation of key epidemiological parameters. An overview is shown in Figure 1.

**Fig. 1:**
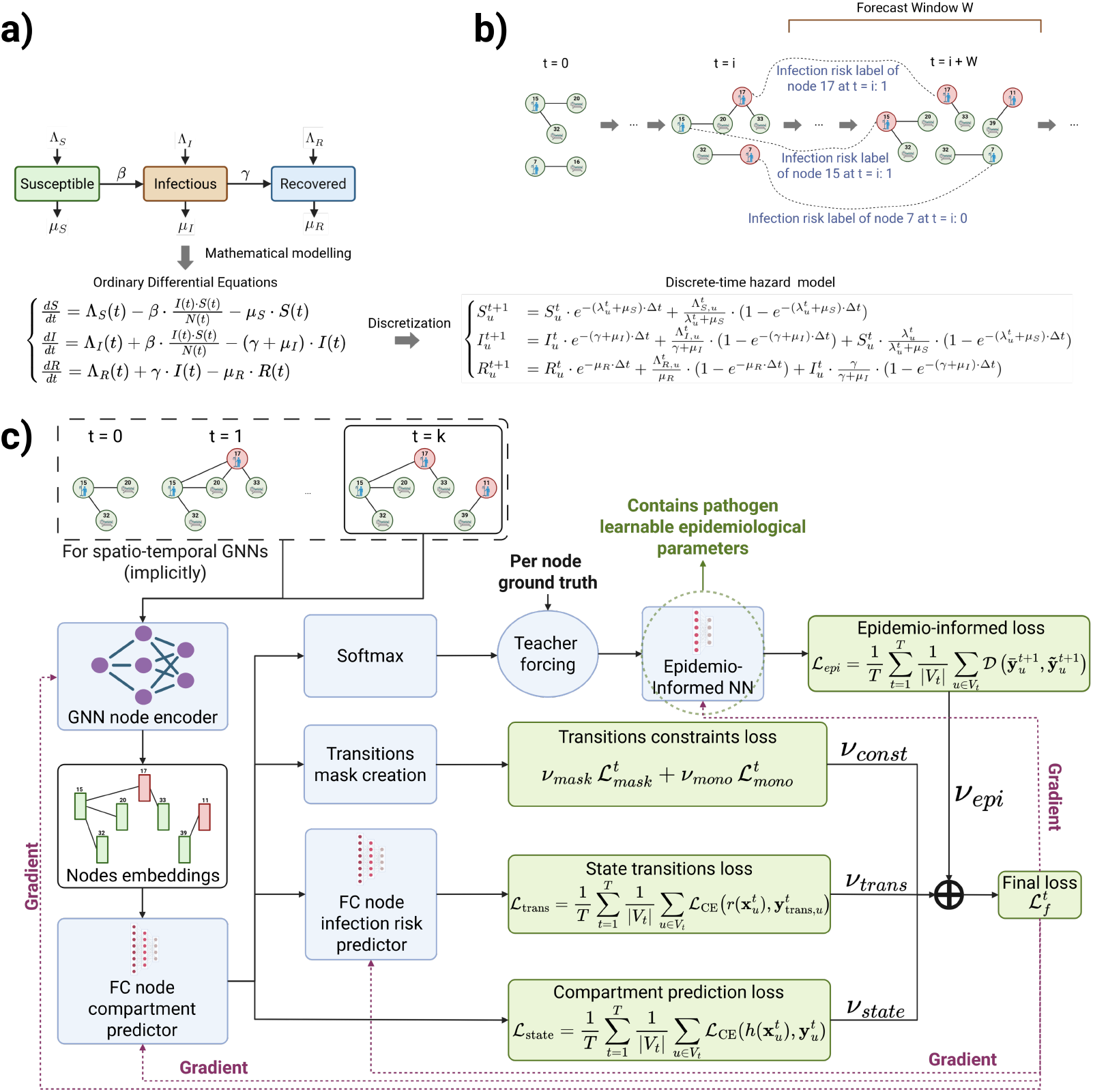
Overview of the proposed EIGNN framework. (a) Mathematical modeling of epidemic dynamics. (b) Temporal sequence graph representation of the hospital state over time. (c) Proposed EIGNN framework. The approach follows a multi-task training paradigm applied to a sequence of temporal graph snapshots representing the hospital contact network. At each time step, a GNN node encoder processes the current graph snapshot, capturing the hospital environment at that moment, and produces node-level embeddings. These embeddings are jointly used to (i) incorporate epidemiology-informed priors through ODE-based regularization (ℒ_*epi*_), (ii) enforce epidemiological state-transition constraints 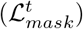, (iii) estimate individual state transitions (ℒ_*trans*_), and (iv) predict individual epidemic states (ℒ_*state*_). Gradients are backpropagated to the GNN encoder, the state transitions prediction head, and the epidemiology-informed neural module, enabling end-to-end optimization and unsupervised learning of epidemiological parameters.

### Predicting Patient-Level States Transitions Performance

We evaluate two epidemiology-informed strategies for infection risk prediction: *AutoDiff*, which enforces consistency of governing ODEs via automatic differentiation, and *ODE-NSP*, which performs next-state prediction using discretized ODEs. Both are compared against non-epidemiology-informed baselines across multiple GNN architectures to assess whether epidemiological constraints improve graph-based models, rather than to benchmark against other temporal modeling paradigms.

Clinical utility is assessed on the real-world *HUG-COVID* dataset [53], capturing COVID-19 infections at the University Hospital of Geneva (2020). Mechanistic consistency is further evaluated on two synthetic datasets: *SocioPatterns* [54, 55], combining real contacts with simulated dynamics, and *Murcia* [56], modeling *Clostridioides difficile* transmission (see Section 5).

Models were evaluated across horizons *W* ∈ {1, 3, 7} days using a standardized classification framework: **Class 0 (Stability)** for stationary states; **Class 1 (Progression)** for active transmission (e.g., *S* → *I*); and **Class 2 (Resolution)** for transitions to absorbing states (e.g., recovery). For the Murcia dataset, a fourth class distinguishes alternative clinical outcomes.

Results for HUG-COVID are shown in Fig. 2 and Appendix 5; SocioPatterns and Murcia results for *W* = 7 are reported in Appendix 5.

**Fig. 2:**
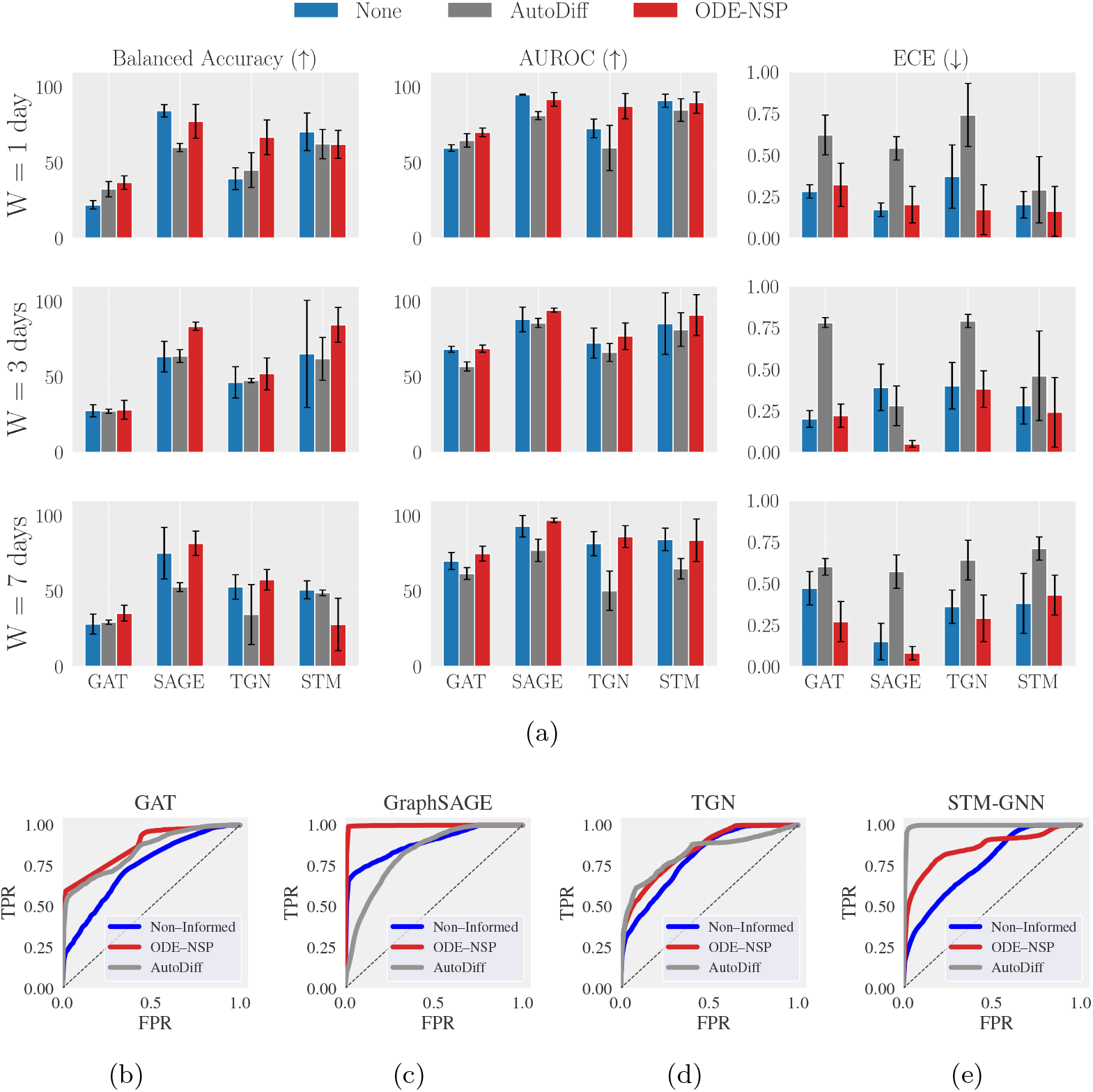
State prediction performance for forecast horizons of 1, 3, and 7 days on the HUG-COVID dataset. (a) Balanced accuracy (first column), AUROC (second column) and ECE (third column) for each forecast horizon (rows). Area under the receiver operating characteristic (AUROC) curve for class 1 (progression), for the different GNN models evaluated on the HUG COVID dataset with a forecast horizon of *W* = 7. Results are shown for (b) GAT, (c) GraphSAGE, (d) TGN, (e) STM-GNN. SAGE and STM refer to GraphSAGE and STM-GNN, respectively.

#### Predictive performance

Models trained with ODE-NSP generally achieve the highest predictive performance on HUG-COVID, followed by non-informed variants, while AutoDiff performs worst. On SocioPatterns and Murcia, performance is more comparable. Additionally, on HUG-COVID, ODE-NSP yields stable results across horizons, reaching 81.53% balanced accuracy and 96.79% AUROC at 7 days, versus 75.01% and 92.84% for non-informed models, showing that the impact of epidemiological regularization is both horizon- and architecture-dependent. At short horizons (1 day), non-informed strong backbones such as GraphSAGE already perform well (balanced accuracy 84.19%, AUC 94.85%), leaving limited room for improvement. However, at longer horizons (3–7 days), epidemiological constraints provide clearer benefits, highlighting that mechanistic priors are particularly valuable when the task shifts from short-term horizons to longer-term ones. These effects also depend on architectural compatibility. Graph-SAGE and TGN often benefit from ODE-NSP, whereas STM-GNN shows mixed behavior, with gains at 3 days (balanced accuracy 65.12% to 84.29%) but degradation at 7 days, suggesting that constraints may become overly restrictive for complex architectures, leading to underfitting or optimization instabilities.

Furthermore, AutoDiff models occasionally match baselines but generally under-perform on HUG-COVID, likely due to optimization complexity and the sensitivity of automatic differentiation to the noise and sparsity characteristic of real-world clinical data. In contrast, ODE-NSP reduces variance (e.g., GraphSAGE at 3 days: 83.35 ± 2.78), indicating improved training stability, while enhancing class separation through simultaneous improvements in sensitivity and specificity. At *W* = 7, STM-GNN with AutoDiff achieves high AUROC for progression (class 1) prediction (99.09%), but performs poorly on stability (class 0) and resolution (class 2) prediction (AUROCs 32.83%, 62.09%), compared with 75.53% and 89.29% for ODE-NSP. This highlights the more balanced performance of ODE-NSP across clinically relevant outcomes, which caputres more reliably persistence and recovery dynamics.

At last, model calibration varies by strategy, with ODE-NSP significantly improving the ECE in HUG-COVID by up to 0.34, whereas AutoDiff often degrades calibration and increases variance. Overall, epidemiology-informed models are most beneficial at intermediate-to-long horizons, where structural priors help stabilize the capacity of the model predict system dynamics beyond the time window directly supported by recent observations. However, gains depend on alignment between mechanistic constraints and architectural design, highlighting the importance of jointly considering forecast horizon, optimization strategy, and model inductive bias when embedding domain knowledge into deep temporal graph models. Importantly, epidemiology-informed models enable the unsupervised learning of parameters governing epidemic dynamics, unlike purely data-driven approaches (see Section 5).

#### Continual model adaptation as a driver of robustness and performance

For a forecast horizon of *W* = 7, we train the different models under a continual learning (CL) paradigm. At each time step *t*, the model is trained using all data available up to and including time *t* (i.e., time steps ≤ *t*), and subsequently evaluated on future observations within the forecast window (e.g., for *W* = 7, evaluation is performed over time steps *t* + 1 to *t* + *W*). Model parameters are not reinitialized between successive time steps; instead, training and evaluation proceed sequentially as time advances. Owing to the substantially higher computational cost of this training procedure, CL models are trained only once, and multiple runs with different random initializations are not performed. The results of this experiment are summarized in Figure 3.

**Fig. 3:**
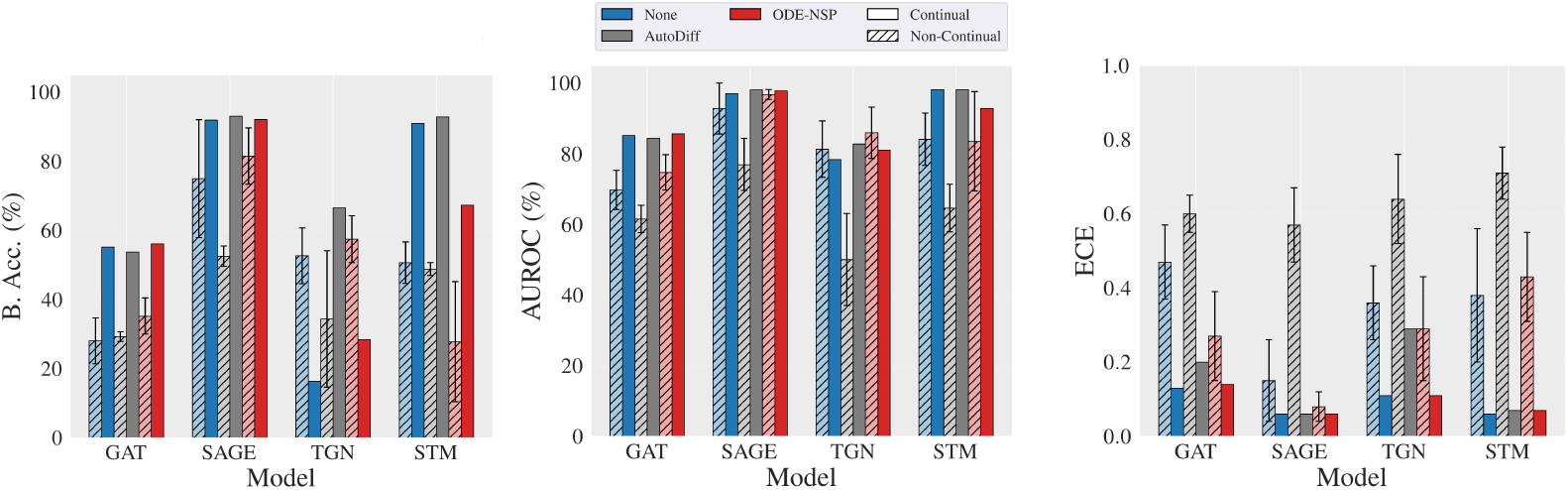
State transition prediction performance metrics for non-continual (striped) vs. continual (unstriped) learning on the HUG-COVID dataset with a 7-day forecast horizon: Balanced accuracy (left), AUROC (middle), ECE (right). SAGE and STM refer to GraphSAGE and STM-GNN, respectively. Error bars indicate standard deviation.

Overall, these results suggest that CL is an important factor for predictive performance under a 7-day forecast horizon. Across most architectures, CL alone (without epidemiological regularization) yields substantial gains over static training. For example, GAT without epidemiological constraints improves from a balanced accuracy of 28.06% to 55.18%, with AUC increasing from 69.77% to 85.23% and ECE decreasing from 0.47 to 0.13. These results indicate that accounting for temporal non-stationarity through sequential adaptation is critical in dynamic hospital epidemic settings, and may be as important as the choice of epidemiological regularization itself.

Importantly, CL largely mitigates the instability previously observed for AutoDiff-based supervision. Under static training, AutoDiff models often exhibited degraded discrimination and poor calibration (e.g., STM-GNN balanced accuracy 48.79%, ECE 0.71). Under CL, the same configuration becomes one of the top-performing models (balanced accuracy 92.89%, AUC 98.21%, ECE 0.07). This result supports the hypothesis that AutoDiff instability stems primarily from static optimization under non-stationary dynamics, rather than from limitations of the epidemiological constraints.

Another important observation is a ceiling effect under CL. For instance, Graph-SAGE achieves balanced accuracies above 92% and ECE values around 0.06 regardless of whether epidemiological regularization is absent, AutoDiff-based, or ODE-NSP-based. This suggests that dynamic updating may implicitly capture part of the evolving epidemic structure, reducing reliance on explicit mechanistic constraints.

Nevertheless, CL does not universally improve performance. For TGN, continual training degrades balanced accuracy in both the non-informed and ODE-NSP settings, indicating that architectures with internal memory mechanisms may not benefit from external sequential training. This highlights the importance of alignment between architectural inductive biases and training strategy.

Finally, CL consistently improves calibration, with ECE values collapsing to the 0.06–0.14 range for most high-performing models. Gains in balanced accuracy are accompanied by simultaneous improvements in sensitivity and specificity, confirming that performance enhancements reflect genuine improvements in class separability rather than threshold adjustments. Overall, these results demonstrate that CL is not only an optimization improvement, but a central component for robust epidemiology-informed prediction in non-stationary clinical environments.

### Unsupervised Learning of Epidemiological Parameters

In this experiment, we investigate a key aspect of our EIGNNs framework: the unsupervised learning of epidemiological parameters. For the HUG-COVID dataset, where the epidemiological parameters are unknown, we evaluate model performance by comparing the global number of patients in each epidemiological state predicted by the model with the corresponding ground-truth counts provided by the data. We compute the daily number of patients in each state using two complementary approaches: (i) direct state prediction with the GraphSAGE AutoDiff model, and (ii) state evolution obtained by applying the discretized ODEs (cf. Section 5) using the epidemiological parameters learned by the GraphSAGE ODE-NSP model.

The comparisons presented in Figure 4 show that both approaches capture temporal trends that closely match the ground truth. These results demonstrate that, although the epidemiological parameters are learned in a fully unsupervised manner, they remain effective for accurately reproducing the global epidemic dynamics.

**Fig. 4:**
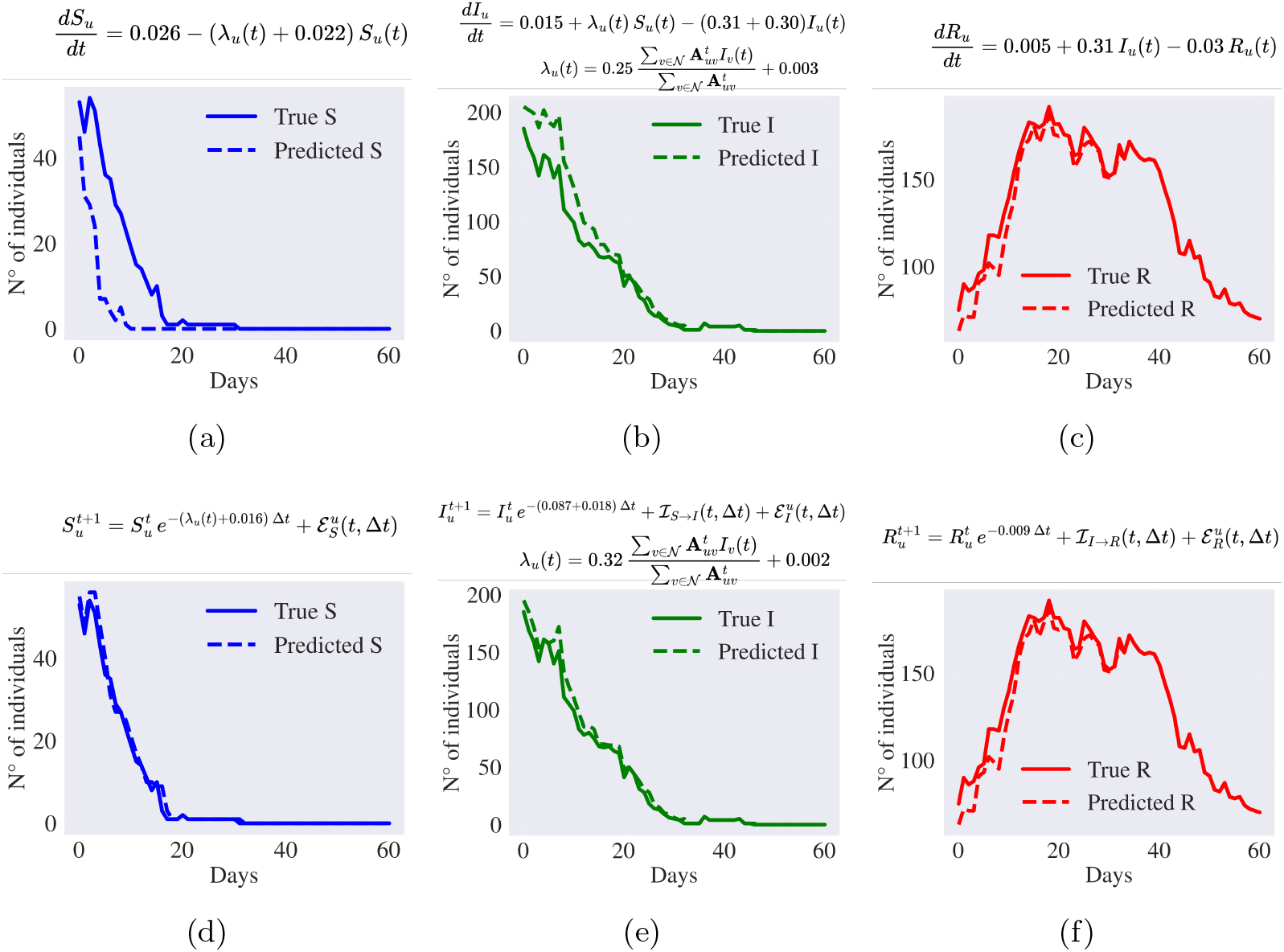
Evolution of the global number of patients in each SIR state over the test period for the HUG-COVID dataset with a forecast horizon of *W* = 7. (a–c) show the results obtained with the GraphSAGE AutoDiff model, while (d–f) correspond to the GraphSAGE ODE-NSP model using discretized equations and learned epidemiological parameters. Solid lines represent the ground-truth number of patients in each state on a given day, whereas dashed lines correspond to the model predictions.

Furthermore, the epidemiological parameters learned by the ODE-NSP model remain biologically plausible. In particular, the estimated recovery period is approximately 12 days, which closely matches the 14-day recovery threshold defined in the HUG-COVID dataset. In addition, with a learned transmission rate of 0.32, the model yields an estimated basic reproduction number of *R*_0_ = 3.68, consistent with reported values in the literature [57, 58].

To further assess the quality of the learned parameters, we also consider the SocioPatterns and Murcia datasets, which are simulated and therefore provide access to the ground-truth epidemiological parameters, assumed to remain constant through-out the simulations. Results for a forecasting horizon of *W* = 7 days are reported in Table 1, while Figure 5 illustrates the per-individual force of infection for the SocioPatterns dataset for the GraphSAGE and STM-GNN models.

**Table 1:**
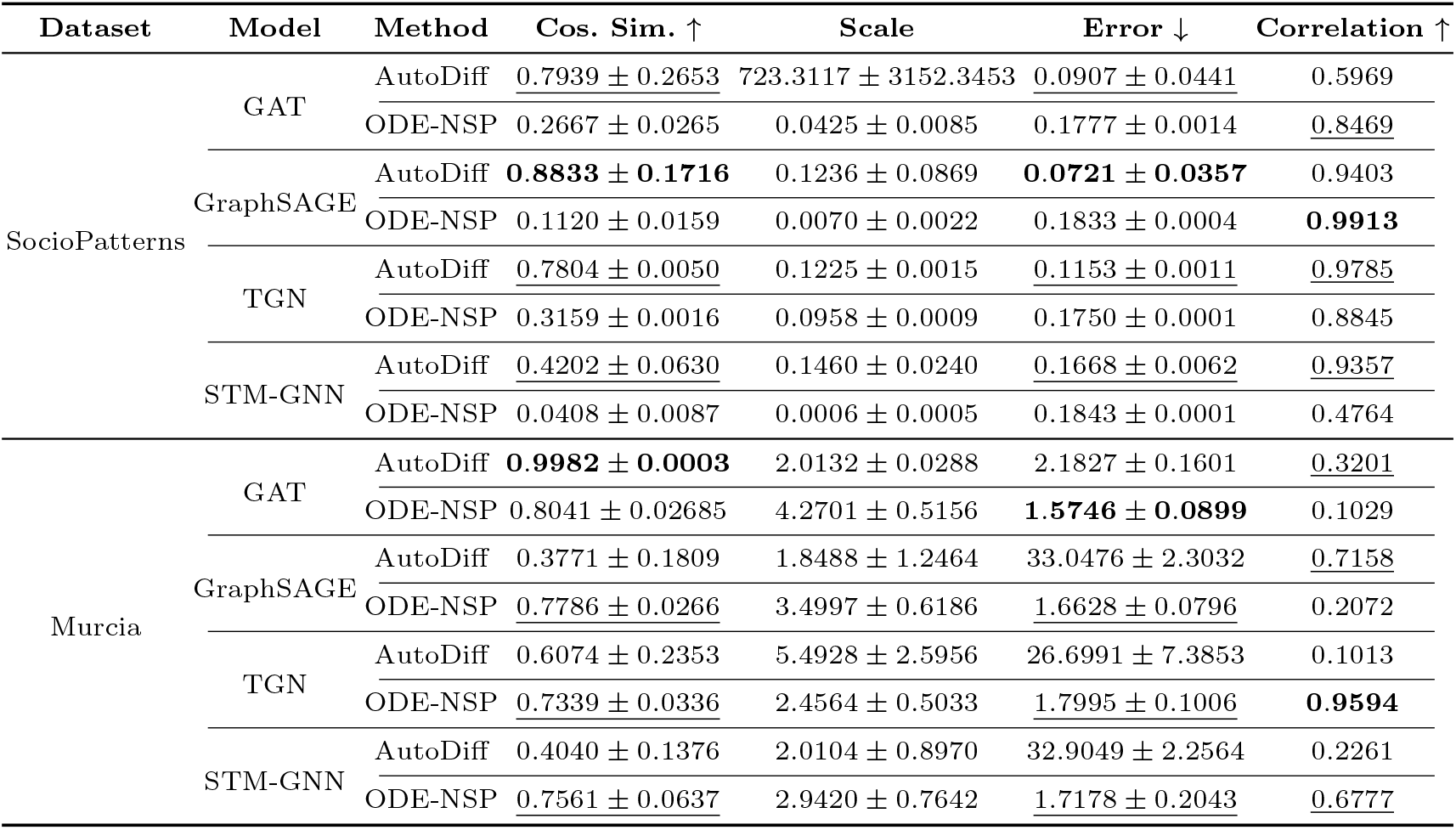
Metrics of the learned epidemiological parameters for a 7-day forecast horizon on the SocioPatterns and Murcia datasets. Method refers to the epidemiology-informed approach used. Scale denotes the optimal scaling factor applied to map the learned parameters to the true parameters. Error represents the norm of the residual error after scaling. Correlation corresponds to the Pearson correlation coefficient of the force of infection.

**Fig. 5:**
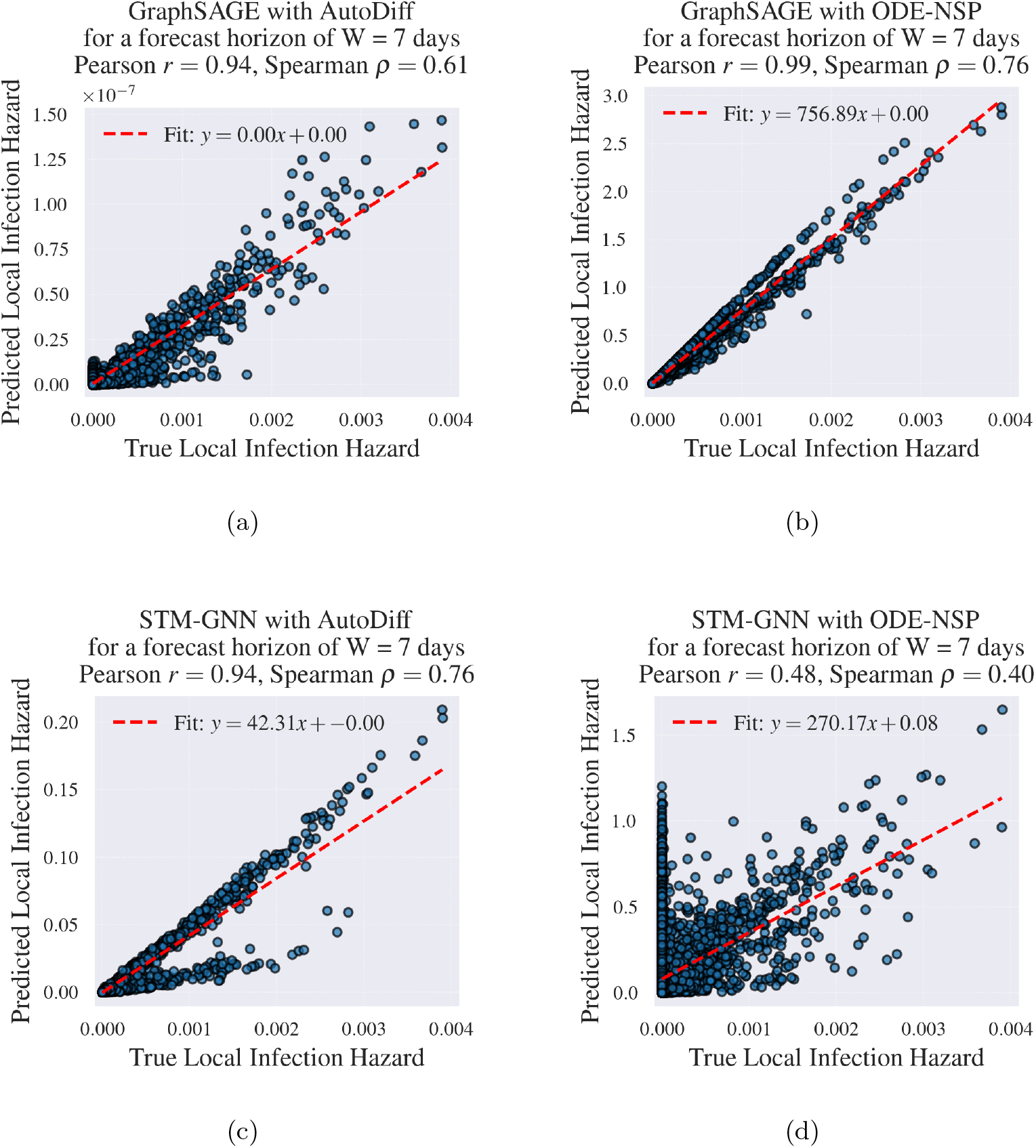
Force of infection estimated for all individuals across all time steps in the SocioPatterns dataset for a forecast horizon of *W* = 7 days. (a) GraphSAGE AutoDiff. (b) GraphSAGE ODE-NSP. (c) STM-GNN AutoDiff. (d) STM-GNN ODE-NSP. Only one model is depicted here, i.e. one that has been trained on a fixed dataset.

Across the SocioPatterns dataset, AutoDiff-based models consistently achieve higher cosine similarity and lower residual error, indicating a closer alignment between the learned and true epidemiological parameters (up to an arbitrary scaling factor). In contrast, ODE-NSP models often achieve higher correlation in the resulting force of infection, suggesting that they reproduce the infection dynamics accurately even when the recovered parameter vectors differ from the ground truth. In addition, the large variability observed in the optimal scaling factors (e.g., 723 ± 3152 for GAT AutoD-iff) further confirms that the epidemiological parameters are identifiable only up to a scaling factor, as expected from the model formulation.

Moreover, the ODE-NSP approach appears to be less stable on the Murcia dataset, where results are often degraded. Overall performance on this dataset is lower and more variable, highlighting its increased complexity. This dataset is larger, contains more fine-grained temporal information, and exhibits noisier dynamics due to its place-based graph modeling of interactions between patients. Specifically, patients located in the same place (e.g., ward, room, etc.) are connected through two-hop paths at a given time, which may link recovered and infected individuals occupying different locations.

Overall, these results highlight a trade-off between accurate recovery of the epidemiological parameters and faithful reproduction of the infection dynamics. While AutoDiff models tend to recover parameter directions more accurately, ODE-NSP models often produce infection dynamics that more closely match the ground truth.

## Discussion

In this work, we introduce a patient-level EIGNN framework designed to provide interpretable infection risk assessments, composed of two methods: AutoDiff, which leverages automatic differentiation to estimate time derivatives of epidemic states and compute residuals from the governing ODEs, and ODE-NSP, which relies on discrete-time approximations of epidemiological ODE models. By integrating mechanistic epidemiological modeling with GNNs, this formulation enables the unsupervised learning of pathogen-specific epidemiological parameters while preserving strong performance in infection-risk prediction. As a result, the proposed models are highly predictive but also more interpretable than purely data-driven approaches, supporting reliable clinical decision-making. Both methods support patient-level infection-risk prediction in hospital contact networks—an application that has received limited attention in prior work. Finally, we evaluated the clinical applicability of our approach on the HUG-COVID cohort and verified its mechanistic fidelity through extensive experiments on two controlled synthetic datasets.

Results on the HUG-COVID dataset highlight the predictive benefits of incorporating epidemiological mechanistic constraints. Overall, ODE-NSP epidemiology-informed models improve both predictive performance and calibration across most architectures and forecast horizons. We observed diverging behaviors in models with strong built-in temporal modeling, such as TGN and STM-GNN. While epidemiological constraints tend to be beneficial for TGN, this is not necessarily the case for STM-GNN, particularly at longer forecast horizons. For STM-GNN, these constraints may partially conflict with the model’s internal temporal representations. This conflict can create optimization trade-offs where the model struggles to reconcile data-driven temporal dependencies with ODE-based mechanistic constraints, potentially leading to underfitting or unstable training as the horizon increases.

More broadly, these results support the interpretation that epidemiological priors act as a *conditional inductive bias*: their utility increases as predictions rely less on immediate contact information and more on latent infection states, incubation periods, and disease progression dynamics. In realistic clinical datasets such as HUG-COVID—characterized by temporal noise, partial observability, and delayed labels—this inductive bias helps regularize the learning process and prevents biologically implausible predictions that may arise in purely data-driven temporal models. By contrast, in highly structured or synthetic environments where contact patterns are cleaner and more deterministic, baseline graph models may already capture most of the predictive signal, thereby reducing the predictive benefit of epidemiological constraints.

Furthermore, we observed an interesting behaviour when CL was introduced: it frequently narrowed the predictive performance gap between AutoDiff and non-informed models relative to ODE-NSP. These findings suggest that the primary benefit of epidemiological regularization lies in its ability to stabilize static training under non-stationary dynamics. However, once continual adaptation is incorporated, the model can directly track evolving epidemic conditions, thereby reducing the reliance on explicit mechanistic constraints.

Overall, our results indicate that robust infection-risk prediction in hospital networks emerges from the interaction of three complementary factors: expressive temporal graph architectures, mechanistic epidemiological priors, and adaptive training strategies capable of handling non-stationary dynamics. While each component individually contributes to performance, their combination provides the most reliable predictions across diverse forecast horizons and data conditions.

Importantly, epidemiology-informed training is not only intended to maximize pre-dictive performance. A key strength of our framework lies in its ability to recover meaningful epidemiological parameters directly from patient contact data—something that has not been explored in previous work on EIGNNs. Our experiments demon-strate that both AutoDiff and ODE-NSP can estimate epidemiological parameters, up to the scaling ambiguities inherent to the ODE formulation. In the case of ODE-NSP, this indeterminacy is closely related to the choice of the time step Δ*t*, since multiplying the epidemiological parameters by a constant factor is equivalent to rescaling the time discretization by the same factor.

Finally, our experiments reveal an interesting distinction between accurate parameter recovery and accurate reconstruction of infection dynamics. Some models achieve relatively poor alignment with the true epidemiological parameters while still producing highly correlated forces of infection, whereas others recover parameter vectors that closely match the ground truth but yield less accurate infection dynamics. These observations indicate that multiple parameter configurations can generate similar epidemic trajectories, reflecting well-known identifiability challenges in epidemiological modeling.

### Clinical implications and translational relevance

From a clinical perspective, our proposed approach provides a framework for individualized and population-level infection risk forecasting that is predictive, interpretable, and actionable. By explicitly embedding epidemiological dynamics into temporal contact graph models, our approach enables clinicians and experts to move beyond short-term, contact-driven alerts toward longer-horizon risk assessments that better reflect disease progression and latent infection states. This capability is particularly relevant in healthcare settings, where delayed detection and intervention can substantially impact patient outcomes and resource utilization.

Importantly, the unsupervised estimation of epidemiological parameters offers a transparent link between model predictions and underlying disease mechanisms, facilitating clinical interpretability and trust. Such parameter estimates may support situational awareness during outbreaks by providing indirect indicators of changing transmission dynamics, even when case labels or ground-truth epidemiological states are sparse or unreliable. Moreover, the improved calibration observed suggests that epidemiology-informed models could support risk-stratified decision-making, such as prioritizing patients for testing, isolation, or enhanced monitoring.

Beyond prediction, the network-based and patient-specific nature of our methods enables translational applications in prospective scenario analysis. By simulating changes in contact patterns or intervention strategies, the framework could inform IPC policies, including patient cohorting, healthcare worker assignment, antibiotic stewardship programs, and social distancing measures within hospitals.

In future work, we plan to further develop EIGNNs for scenario modeling and to extend the framework to more advanced CL settings tailored for long-term epidemic monitoring. Such settings would enable continuous model updating as new data become available, without expensive computational costs. Additionally, we aim to explore uncertainty-aware extensions, in which predictions are expressed as probability distributions, providing clinicians and decision-makers with explicit confidence estimates alongside infection risk predictions.

### Limitations

First, although we evaluated the proposed framework on three different datasets, only one corresponds to real-world data (HUG-COVID), while the other two are pseudo-synthetic (SocioPatterns) or fully synthetic (Murcia). The HUG-COVID dataset, in particular, is limited by the relatively small amount of available patient-level information, such as age, sex, or comorbidities, which could be highly informative for predicting individual infection risk. In addition, the observation period covered by this dataset is relatively short (approximately four months), which may limit the ability to capture longer-term epidemic dynamics.

Second, the evaluation of the learned epidemiological parameters is not straight-forward. This issue is especially pronounced for the Murcia dataset, for which only estimated rather than ground-truth epidemiological parameters are available, due to the stochastic nature of the simulation process used to generate the data. As a result, discrepancies between inferred and true parameters may reflect simulation variability rather than model inaccuracies.

Finally, the proposed epidemiology-informed models are inherently sensitive to the validity of the underlying ODE formulations and their discretization. If there is a substantial mismatch between the epidemiological model assumptions and the true data-generating process, or when discretization errors introduce significant approximation bias, the performance of the EIGNNs framework may be adversely affected.

## Methods

### Problem formulation

We formulate the infection risk prediction task as a state transitions and node classification problem on dynamic graphs. Let 𝒢 = {*G*_1_, *G*_2_, …, *G*_*T*_ } denote a sequence of graphs evolving over time steps *t* = 1, …, *T* . Each graph *G*_*t*_ = (*V*_*t*_, *E*_*t*_) consists of a set of nodes *V*_*t*_ (representing patients and/or healthcare workers) and a set of edges *E*_*t*_ representing their interactions. Each node *u* ∈ *V*_*t*_ is associated with a feature vector 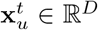 and a corresponding label 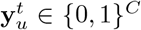 (e.g., patient state), represented as a one-hot encoding over *C* classes.

Our objective is to learn a mapping function *h* that predicts the class probability vector for each node based on its features and network position. We decompose *h* into two components: a node embedding function (encoder) *g* : ℝ^*D*^ → ℝ^*d*^ that maps high-dimensional input features to a latent space, and a classifier *f* : ℝ^*d*^ → [0, 1]^*C*^ that maps these embeddings to probability scores, such that *h* = *f* ◦*g*. The model is trained by minimizing a classification loss function ℒ (e.g., Cross Entropy) via empirical risk minimization:

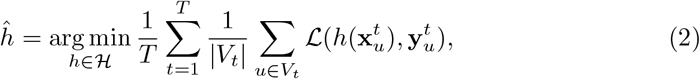

where |*V*_*t*_| denotes the number of nodes at time *t*. For notation simplicity, we describe the general framework for homogeneous graphs. However, the method naturally extends to heterogeneous settings by using type-specific projection matrices in the encoder *g*.

### Datasets

We evaluate our framework on three distinct datasets ranging from real-world hospital data to controlled synthetic environments. The availability of large-scale patient-level contact datasets with reliable infection labels is limited; the HUG-COVID dataset [53] represents one of the few documented real-world hospital contact networks. The synthetic and pseudo-synthetic datasets complement the real data by enabling controlled analysis of model behavior under known regimes.

#### HUG-COVID (Real-World)

This dataset is a subset of the data collected at the *Hôpitaux Universitaires de Genève* (HUG) during the COVID-19 pandemic [53]. It includes 282 hospitalized patients (52 nosocomial cases) observed between 2020-01-21 and 2020-06-03 (135 days).

##### Graph construction

We model the data as a sequence of daily, bidirectional heterogeneous graphs, as illustrated in Fig. 1.b. Nodes represent patients and wards. An edge connects a patient to a ward if the patient was present in that ward on the corresponding day; all edges are assigned unit weight.

##### Node features

Patient nodes are characterized by four features: (i) current infection status (binary), (ii) current epidemiological SIR state (one-hot encoded), (iii) colonization pressure [7], and (iv) time duration in the current SIR state. Ward nodes are characterized by two features: (i) the number of patients present and (ii) ward-level colonization pressure.

##### Prediction task

We define three forecast horizons, *W* ∈ {1, 3, 7} days, where each sample at time *t* is assigned to a class based on state transitions occurring within the window [*t, t*+*W* ]. Specifically, **Class 0 (Stability)** denotes the absence of transitions, **Class 1 (Progression)** encompasses disease evolution such as *S* → *I* or *S* → *I* → *R*, and **Class 2 (Resolution/Other)** accounts for clinical recovery (e.g., *I* → *R*) or transitions not explicitly captured by the epidemiological model. Additionally, the dataset is split temporally into a training set (first 78 days), including 258 patients, and a test set (remaining 57 days), including 213 patients. Detailed statistics for each split are provided in Appendix 5. Feature normalization using a standard scaler for the number of patients per ward and a min-max scaler for the timestamps of each snapshot is computed exclusively on the training set to prevent information leakage.

#### SocioPatterns (Pseudo-Synthetic)

This dataset is a pseudo-synthetic dataset derived from two publicly available contact-network datasets: the SFHH conference dataset [54] and the Lyon hospital ward dataset [55]. Both datasets capture face-to-face proximity interactions using wearable sensors. The SFHH dataset tracks 403 participants over a two-day scientific conference, whereas the Lyon hospital ward dataset follows 75 individuals (29 patients and 46 healthcare workers) over a five-day period. In both cases, the sensors record the cumulative duration of close-range interactions between pairs of individuals.

##### Simulation

Based on these empirical contact networks, we generated 20 independent pseudo-synthetic datasets, following a SEIR model, each spanning between 50 and 200 days. These datasets preserve the statistical properties of the original interaction patterns while enabling controlled epidemiological simulations over extended time horizons. Details regarding dataset generation, graph construction, pre-processing, and data splitting are provided in Appendix 5.

#### Murcia (Synthetic)

The Murcia dataset is a fully simulated dataset generated using the spatiotemporal epidemic simulator introduced in [56]. The simulator models a SEIRD–NS epidemiological process within a hospital environment at a temporal resolution of 8 hours, corresponding to healthcare worker shift changes.

##### Simulation

Using a fixed hospital layout, we generated 10 independent datasets, each covering approximately 2 years. Across simulations, it results in around 26,628 patients distributed across 236 beds and 102 rooms, including emergency rooms, intensive care units, and operating rooms, among others. Additional details on graph modeling, pre-processing, and data splitting procedures are provided in Appendix 5.

### Epidemiological Models

We consider three compartmental epidemiological models—SIR with immigration/emigration, SEIR, and SEIRD-NS—describing the dynamics of the evolution of an epidemic event. In these models, each node u at time t is associated with a probability distribution over epidemiological states (e.g., susceptible *S*_*u*_(*t*), infected *I*_*u*_(*t*), recovered *R*_*u*_(*t*)), summing to 1.

#### Continuous Network-Based SIR Model with Immigration/Emigration

The continuous time-evolution of the infection is governed by a set of ODEs. For the SIR model with open-population dynamics (immigration and emigration), the governing equations for node *u* are:

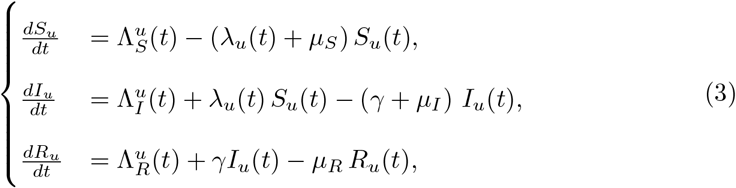

where:

- 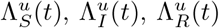 represent the time-dependent external arrival rates (immigration) into each compartment for node *u*.
- *µ*_*S*_, *µ*_*I*_, *µ*_*R*_ are the state-specific departure rates (emigration, transfer, discharge, or death).
- *γ* is the recovery rate.
- *λ*_*u*_(*t*) is the node-specific force of infection, defined as:

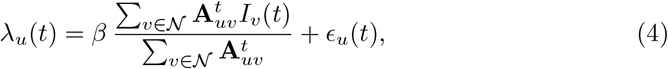

where *β* is the transmission rate, 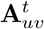 represents the contact weight (e.g., duration if available, 1 otherwise) between node *u* and neighbor *v*, and *ϵ*_*u*_(*t*) accounts for background environmental risk (seeding).

#### Discrete-time hazard approximation

We can discretize the ODEs described in Eq. 3 using an exponential waiting-time framework (Poisson process). Unlike simple Euler discretization, this approach respects the stochastic nature of transitions and competing risks (e.g., a node leaving the system vs. becoming infected).

We assume that the transition rates (such as the force of infection *λ*_*u*_(*t*) or the state-specific arrival rates 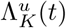) remain constant over the small interval [*t, t* + Δ*t*]. Under this assumption, the state update equations for node *u* are derived using competing-risks theory as follows:

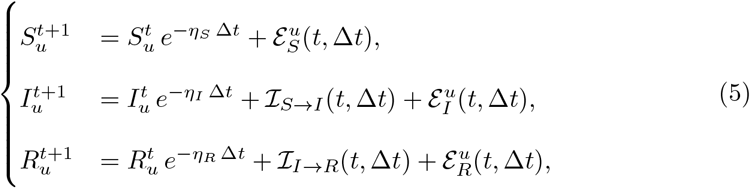

where:

- 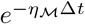 represents the survival probability of state ℳ (staying in the current state), where *η*_*S*_ = *λ*_*u*_(*t*) + *µ*_*S*_, *η*_*I*_ = *γ* + *µ*_*I*_, and *η*_*R*_ = *µ*_*R*_.
- 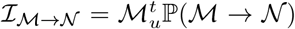 represents the internal inflow from state ℳ to state 𝒩 derived from competing risks (e.g., infection vs. emigration).
- 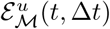 represents the external inflow (i.e. cumulative immigration) into state ℳ over the interval Δ*t*.

In all our experiments, we assume arrival and departure rates 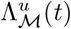 and *µ*_ℳ_ constant for any state ℳ. Detailed approximations of the internal and external inflows for each state can be found in Appendix 5. At last, the formulation for the SEIR and SEIRD-NS models are presented in Appendix 5.

### Network-based Epidemiology-Informed Framework

We propose two distinct strategies to embed the epidemiological dynamics into the GNN training process. Both strategies allow for the unsupervised learning of epidemiological parameters (e.g., *β, γ*) by enforcing consistency between the data-driven predictions and the governing physical laws.

Common to both methods is the computation of the force of infection *λ*_*u*_(*t*). We assume this force is driven by the dynamic contact network encoded in the adjacency matrix **A**^*t*^. As interactions evolve, the GNN leverages the learned node embeddings and the graph structure to estimate the instantaneous risk *λ*_*u*_(*t*) for every individual, which then drives the state transitions in the following modules.

#### Method 1: Continuous-Time Residual Minimization (AutoDiff)

This method enforces the continuous-time dynamics directly using Automatic Differentiation (AutoDiff), analogous to the standard PINNs framework:

1. Time-Aware Embeddings: We augment the node embeddings with an explicit time variable, **u** = [*g*(**x**), *t*], allowing the network to learn continuous temporal functions.
2. Residual Computation: We compute the time derivative of the next state, 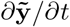, either by using automatic differentiation (i.e., 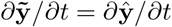) or by discretization based on the true next state (i.e., 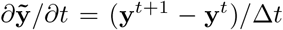). Simultaneously, we compute the expected derivative, 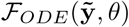, by plugging either the predicted (i.e., 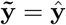) or true states 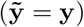 into the governing ODEs defined in Section 5 (Eq. 3).
3. Loss Function: The loss minimizes the distance between the neural derivative and the physical derivative, using a distance function 𝒟:

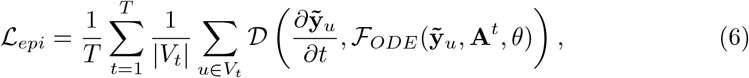

This ensures that the learned trajectory satisfies the fundamental differential equations of the epidemiological model, while learning the epidemiological parameters *θ* in an unsupervised manner.

#### Method 2: Discrete-Time Next State Prediction (ODE-NSP)

Alternatively, we treat the discrete-time update equations (defined in Section 5, Eq. 1) as a differentiable physics layer *p*_*θ*_ parameterized by the learnable epidemiological parameters *θ* = {*β, γ*, Λ_*S*_, Λ_*I*_, Λ_*R*_, *µ*_*S*_, *µ*_*I*_, *µ*_*R*_} .

The physics layer *p*_*θ*_ takes this state and the network structure as input and outputs the expected state at the next time step according to the hazard equations (Eq. 1):

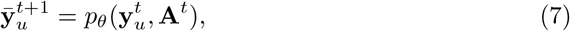

We then minimize the discrepancy between this physics-based prediction, 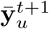, and the target next state (either the ground-truth label, 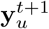, or the model’s own prediction, 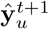) using a regularization loss:

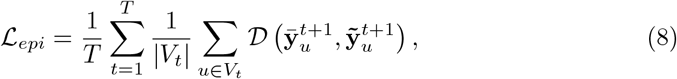

where 𝒟 is a distance metric (e.g., MSE or Cross-Entropy), and 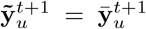 or 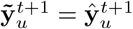, based on the teacher forcing training strategy (see Section 5).

### Handling Large Forecast Horizons

#### One-shot states forecast prediction

Let *W* denote the length of the forecast horizon, and let *C* be the number of states in the epidemiological model under consideration. At a given time step *t >* 0, our goal is to predict the future states for all *k* ∈ [*t* + 1, *t* + *W* ], using as input the feature vectors of the patients/individuals (nodes) at time *t*, denoted by 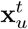, where *u* ∈ [1, |*V*_*t*_|].

In this setting, we adapt our original problem so that the model outputs a sequence of predictions over the horizon instead of a single prediction:

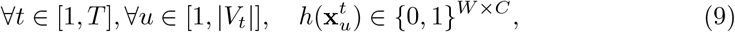

Here, *h* is designed to output, for each sample, a matrix of dimension *W* ×*C*, where the entry 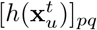 represents the probability that patient/individual *u* is in state *q* at time step *t* + *p*.

#### Guided Training for State Transition Prediction

A primary clinical objective in epidemiological modeling is the characterization of state transitions—specifically, predicting whether an individual will progress from their current health state to another one over the forecast horizon. To do this, we frame the transition dynamics as a multi-class classification problem, categorizing outcomes into distinct trajectories: stability (no transition), infection progression (transitions involving the infectious state), recovery (transitions from I to R), and alternative outcomes (including transitions to absorbing states or non-infection-related paths). In the HUG-COVID and SocioPatterns datasets, recovery and alternative outcomes are grouped together in one class, whereas in the and Murcia dataset, all four classes are used.

Furthermore, within our epidemiology-informed framework, the predicted state distribution is an essential component for enforcing physical consistency via the epidemiological loss ℒ_epi_ (Eq. 6 and Eq. 8). To maintain this constraint, the model architecture outputs the full state distribution. Simultaneously, the underlying node embeddings are used by an auxiliary model *r* (e.g., a MLP) to predict state transitions:

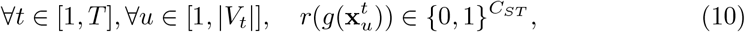

or, for the multi-step case,

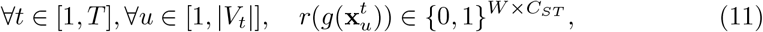

where *C*_*ST*_ is the number of predefined state transitions (*C*_*ST*_ = 3 for HUG-COVID and SocioPatterns, and *C*_*ST*_ = 4 for Murcia).

This shared node embedding space ensures that transition dynamics remain intrinsically coupled with the predicted epidemiological states, and allows us to predict state transitions while guiding training through the state predictions, using the following combined loss:

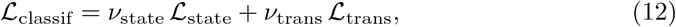

with,

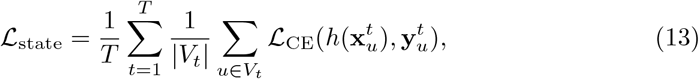

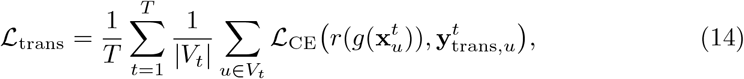

where 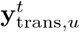 is the multi-class state transition label for patient/individual *u* at time step *t*.

The final prediction of the model is given by *r* and optimized through ℒ_trans_. The additional state-based term ℒ_state_ serves as a training regularizer, encouraging the model to also learn future state dynamics—an aspect particularly valuable in epidemiology-informed settings.

#### Transitions Constraints Loss

##### Masked Loss

Based on the epidemiological parameters and the temporal resolution of our dataset (e.g., daily snapshots), certain state transitions are biologically infeasible within a single time step. For instance, given typical incubation and recovery periods, a patient cannot transition directly from a Susceptible state at day *t* to a Recovered state at day *t* + 1 without first progressing through an Infectious stage.

To enforce these physical constraints across the forecast window *W*, we propose a transition constraint loss ℒ_*mask*_ that penalizes the model for assigning probability mass to unreachable states. Let 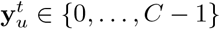 denote the ground truth state of patient *u* at step *t* within the forecast window. We define a transition compatibility matrix 𝒯 ∈ {0, 1}^*C*×*C*^, where 𝒯_*i,j*_ = 1 if a transition from state *i* to state *j* is valid, and 𝒯_*i,j*_ = 0 otherwise.

For each patient *u* and each step *t*, we identify forbidden transitions within the forecast window based on the observed state at the previous step. The masked loss is defined as:

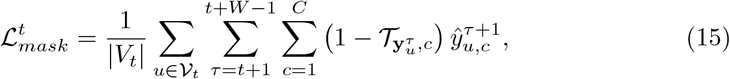

where 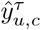 denotes the predicted probability of patient *u* being in state *c* at forecast step *τ* . For new patients appearing at time *τ* +1, we assume their initial state is always valid; thus, they are naturally handled by the initialization of the temporal graph.

##### Monotonicity Loss

To further enforce temporal and self-consistency in the forecast window *W*, we propose a monotonicity loss 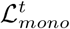. We compute the expected predicted state score, 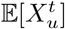, as the weighted average of the probabilities across all potential states for patient *u* at time *t*. Using this, we define a differentiable constraint that penalizes deviations from the expected disease progression trajectory, specifically targeting instances where the expected state score increases over time:

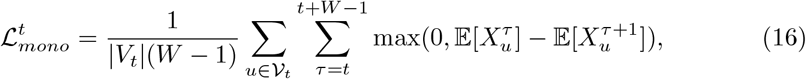

where 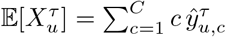 is the expected predicted state score.

This loss funciton allows to penalize biologically implausible transitions (e.g., I→S) within the prediction window without requiring explicit rule-based post-processing.

##### Final Constraints Loss

The final transitions constraints loss function is a weighted sum of the previous two losses (15) and (16):

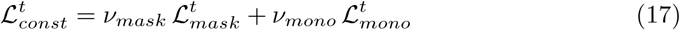

### Final Multi-Task Epidemiology-Informed Loss Function

The final classification loss function to optimize is a weighted combination of the loss functions ℒ_*trans*_ and ℒ_*state*_ from Eq. (12), the epidemiology-informed loss function ℒ_*epi*_ from Eqs. (6) and ( 8), and the transitions constraints loss function 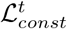 from Eq. (17) where the corresponding weights are denoted by *ν*_*trans*_, *ν*_*state*_, *ν*_*epi*_, and *ν*_*mask*_ respectively:

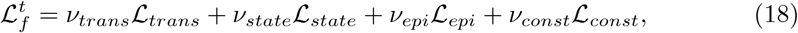

### Teacher Forcing Strategy

To ensure a more stable training process in the early stages, we do not solely rely on the predicted future states of the individuals/nodes obtained through *h*, but instead also use the true next states (i.e., the labels), following a teacher forcing strategy. Specifically, we use the true labels with a probability *p*_*T F*_ and the predicted labels with a probability 1 − *p*_*T F*_ . In addition, the weight of the epidemiology-informed loss *ν*_*epi*_ is initially set to a small value (giving more importance to the classification loss) and is progressively increased during the training.

To this end, we define a simple hybrid teacher forcing scheduling strategy in which:

- *ν*_*epi*_ increases linearly over the first *ramp* epochs and then remains constant at a predefined value 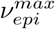:
- teacher forcing is applied with a probability 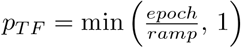.

At the beginning of the training, the model updates mainly rely on teacher forcing. Once the *ramp* epochs have passed, the model’s own predictions are increasingly used to compute the different terms of the ODEs.

### Experimental Setup

We conducted a series of experiments to validate and evaluate our network-based EIGNNs framework. To this end, we considered multiple GNN architectures for the node encoder *g*, including GAT [59], GraphSAGE [60], TGN [18], and STM-GNN [8]. For the predictor *f*, we employed a linear layer followed by either a sigmoid activation function for binary classification or a softmax activation function for multi-class classification. Additionally, the weights of all models were randomly initialized, with the exception of the epidemic models *p*_*θ*_ and F_ODE_. The weights for these specific models were initialized using plausible epidemiological parameter values to mitigate arbitrary scaling and translation of the learned parameters. Detailed initialization procedures are provided in Appendix 5.

Moreover, models were trained—across all forecast horizons—to jointly predict state transitions and epidemiological state. Across all experiments, model parameters were optimized using the Adam optimizer with a weighted cross-entropy loss to address class imbalance for the prediction tasks, and a mean squared error (MSE) loss for the epidemiology-informed terms. Models were trained for up to 50 epochs with a weight decay of 1 × 10^−7^. Early stopping was applied with a patience of 15 epochs and a minimum improvement threshold of 1 × 10^−5^. For the STM-GNN model, the memory size was fixed to 10 for all the datasets. All remaining hyperparameters were optimized using Optuna [61], with the objective of maximizing AUC ROC over the state transitions. The models and hyperparameters used for training are shared with the code.

### Evaluation Framework

All experiments were repeated multiple times to account for stochastic variability. Specifically, variability was introduced either by using different independently generated datasets (SocioPatterns and Murcia), or by using different random weight initializations on the same dataset (HUG COVID). Reported results therefore reflect averaged performance across runs, providing a more robust assessment of model stability and generalization.

For model evaluation, we report standard classification metrics including sensitivity, specificity, adjusted balanced accuracy (between −1 and 1), area under the receiver operating characteristic (ROC) curve (AUC), and expected calibration error (ECE). These metrics were selected to jointly assess discrimination, class-wise performance under imbalance, and probabilistic calibration.

To evaluate the epidemiological parameters learned in an unsupervised manner, we employ metrics that explicitly account for the inherent indeterminacy in scale and translation. All learned epidemiological parameters are concatenated into a single vector and compared to the corresponding vector of ground-truth parameters. We report the cosine similarity, which is invariant to scaling, as well as the residual error after optimal scaling. In addition, we compute the Pearson correlation coefficient (PCC) between the true and learned patient-level forces of infection *λ*_*u*_ (as defined in Section 5).

## Data Availability

Data from the HUG cohort are subject to institutional privacy restrictions and cannot be publicly shared. All other data supporting the findings of this study are available from the corresponding author upon reasonable request.

## Code availability

The source code for this EIGNN framework, as presented in this study, is publicly available at https://github.com/yamilvindas/eignnshai.

## Acknowledgements

We thank Prof. José M. Juarez and Dr. Denisse Kim for helpful discussions and for clarifying details of their *Clostridioides difficile* HAI simulator developed by their team, which greatly aided our understanding of the data.

## Declarations

### Ethics approval and consent to participate

The analysis related to the HUG dataset was approved by the Geneva Cantonal Ethics Committee [number CCER 2020–00827 15\LO\0746].

Editorial Policies for:

Springer journals and proceedings: https://www.springer.com/gp/editorial-policies

Nature Portfolio journals: https://www.nature.com/nature-research/editorial-policies

*Scientific Reports*: https://www.nature.com/srep/journal-policies/editorial-policies

BMC journals: https://www.biomedcentral.com/getpublished/editorial-policies

## State Transition Prediction for Forecasts Horizons of 1, 3 and 7 Days on the HUG-COVID dataset

**Table 1:** State transition prediction performance for forecast horizons of 1, 3, and 7 days on the HUG-COVID dataset.

| Forecast horizon (days) | Model | Epidemiology-informed method | Sensitivity ↑ | Specificity ↑ | Balanced acc. ↑ | AUC ↑ | ECE ↓ |
| --- | --- | --- | --- | --- | --- | --- | --- |
| 1 | GAT | None | $47.97 \pm 1.80$ | $68.04 \pm 0.39$ | $21.96 \pm 2.71$ | $59.52 \pm 2.25$ | $0.28 \pm 0.04$ |
| | | AutoDiff | $54.89 \pm 3.37$ | $68.58 \pm 1.83$ | $32.34 \pm 5.06$ | $64.67 \pm 4.49$ | $0.62 \pm 0.12$ |
| | | ODE-NSP | <u>$57.77 \pm 2.98$</u> | <u>$71.29 \pm 3.12$</u> | <u>$36.66 \pm 4.48$</u> | <u>$69.96 \pm 2.91$</u> | $0.32 \pm 0.13$ |
| | GraphSAGE | None | **$89.46 \pm 2.73$** | **$89.60 \pm 2.73$** | **$84.19 \pm 4.09$** | **$94.85 \pm 0.39$** | $0.17 \pm 0.04$ |
| | | AutoDiff | $73.28 \pm 1.88$ | $73.86 \pm 1.84$ | $59.93 \pm 2.82$ | $80.96 \pm 2.67$ | $0.54 \pm 0.07$ |
| | | ODE-NSP | $84.86 \pm 7.52$ | $86.68 \pm 6.47$ | $77.28 \pm 11.27$ | $91.78 \pm 4.59$ | $0.20 \pm 0.11$ |
| | TGN | None | $59.50 \pm 4.78$ | $71.76 \pm 4.47$ | $39.24 \pm 7.17$ | $72.55 \pm 6.22$ | $0.37 \pm 0.19$ |
| | | AutoDiff | $63.28 \pm 7.68$ | $68.33 \pm 5.34$ | $44.92 \pm 11.51$ | $59.63 \pm 14.92$ | $0.74 \pm 0.19$ |
| | | ODE-NSP | <u>$77.79 \pm 7.70$</u> | <u>$83.52 \pm 6.72$</u> | <u>$66.69 \pm 11.55$</u> | <u>$87.32 \pm 8.37$</u> | $0.17 \pm 0.15$ |
| | STM-GNN | None | <u>$80.18 \pm 8.33$</u> | <u>$87.82 \pm 4.34$</u> | <u>$70.28 \pm 12.49$</u> | <u>$90.90 \pm 4.33$</u> | $0.20 \pm 0.08$ |
| | | AutoDiff | $74.83 \pm 6.43$ | $81.77 \pm 7.44$ | $62.24 \pm 9.65$ | $84.73 \pm 7.53$ | $0.29 \pm 0.20$ |
| | | ODE-NSP | $74.60 \pm 6.18$ | $86.79 \pm 6.30$ | $61.90 \pm 9.27$ | $89.73 \pm 7.03$ | **$0.16 \pm 0.15$** |
| 3 | GAT | None | $51.64 \pm 2.67$ | $70.68 \pm 2.13$ | $27.47 \pm 4.01$ | $68.11 \pm 1.98$ | <u>$0.20 \pm 0.05$</u> |
| | | AutoDiff | $51.40 \pm 0.94$ | $67.53 \pm 0.12$ | $27.10 \pm 1.40$ | $56.75 \pm 2.96$ | $0.78 \pm 0.03$ |
| | | ODE-NSP | <u>$52.06 \pm 4.12$</u> | <u>$71.73 \pm 2.46$</u> | <u>$28.09 \pm 6.17$</u> | <u>$68.66 \pm 2.40$</u> | $0.22 \pm 0.07$ |
| | GraphSAGE | None | $75.58 \pm 6.76$ | $77.83 \pm 6.18$ | $63.37 \pm 10.14$ | $87.90 \pm 8.18$ | $0.39 \pm 0.14$ |
| | | AutoDiff | $75.78 \pm 2.85$ | $78.82 \pm 3.39$ | $63.67 \pm 4.28$ | $85.61 \pm 2.97$ | $0.28 \pm 0.12$ |
|  |  | ODE-NSP | <u>$88.90 \pm 1.85$</u> | **$90.50 \pm 1.59$** | <u>$83.35 \pm 2.78$</u> | **$94.04 \pm 1.31$** | **$0.05 \pm 0.02$** |
| | TGN | None | $64.20 \pm 6.89$ | $72.20 \pm 4.14$ | $46.29 \pm 10.33$ | $72.25 \pm 9.96$ | $0.40 \pm 0.14$ |
| | | AutoDiff | $64.94 \pm 0.94$ | $68.40 \pm 0.20$ | $47.41 \pm 1.40$ | $66.16 \pm 5.97$ | $0.79 \pm 0.04$ |
|  |  | ODE-NSP | <u>$67.92 \pm 7.04$</u> | <u>$72.92 \pm 6.11$</u> | <u>$51.88 \pm 10.56$</u> | <u>$76.78 \pm 8.81$</u> | <u>$0.38 \pm 0.11$</u> |
| | STM-GNN | None | $76.75 \pm 23.62$ | $86.84 \pm 10.19$ | $65.12 \pm 35.43$ | $85.11 \pm 20.44$ | $0.28 \pm 0.11$ |
| | | AutoDiff | $74.58 \pm 9.42$ | $75.24 \pm 9.25$ | $61.86 \pm 14.13$ | $81.15 \pm 11.12$ | $0.46 \pm 0.27$ |
|  |  | ODE-NSP | **$89.53 \pm 7.69$** | <u>$90.13 \pm 7.62$</u> | **$84.29 \pm 11.54$** | <u>$90.70 \pm 13.53$</u> | <u>$0.24 \pm 0.21$</u> |
| 7 | GAT | None | $52.04 \pm 4.41$ | $70.31 \pm 2.98$ | $28.06 \pm 6.62$ | $69.77 \pm 5.56$ | $0.47 \pm 0.10$ |
| | | AutoDiff | $52.85 \pm 0.94$ | $68.34 \pm 0.70$ | $29.28 \pm 1.42$ | $61.52 \pm 3.86$ | $0.60 \pm 0.05$ |
|  |  | ODE-NSP | <u>$56.81 \pm 3.48$</u> | <u>$74.34 \pm 3.26$</u> | <u>$35.21 \pm 5.22$</u> | <u>$74.76 \pm 4.98$</u> | <u>$0.27 \pm 0.12$</u> |
| | GraphSAGE | None | $83.34 \pm 11.38$ | $87.57 \pm 6.93$ | $75.01 \pm 17.07$ | $92.84 \pm 7.21$ | $0.15 \pm 0.11$ |
| | | AutoDiff | $68.36 \pm 1.93$ | $72.76 \pm 1.60$ | $52.54 \pm 2.90$ | $76.96 \pm 7.37$ | $0.57 \pm 0.10$ |
|  |  | ODE-NSP | **$87.69 \pm 5.41$** | **$90.57 \pm 4.02$** | **$81.53 \pm 8.12$** | **$96.79 \pm 1.43$** | **$0.08 \pm 0.04$** |
| | TGN | None | $68.43 \pm 5.39$ | $75.36 \pm 3.79$ | $52.64 \pm 8.09$ | $81.31 \pm 7.98$ | $0.36 \pm 0.10$ |
| | | AutoDiff | $56.23 \pm 13.19$ | $67.40 \pm 1.64$ | $34.35 \pm 19.78$ | $50.08 \pm 13.03$ | $0.64 \pm 0.12$ |
|  |  | ODE-NSP | <u>$71.62 \pm 4.53$</u> | <u>$77.17 \pm 3.78$</u> | <u>$57.44 \pm 6.79$</u> | <u>$85.98 \pm 7.27$</u> | <u>$0.29 \pm 0.14$</u> |
|  | STM-GNN | None | <u>$67.12 \pm 3.99$</u> | <u>$77.90 \pm 5.39$</u> | <u>$50.68 \pm 5.99$</u> | <u>$84.11 \pm 7.42$</u> | <u>$0.38 \pm 0.18$</u> |
| | | AutoDiff | $65.86 \pm 1.24$ | $68.25 \pm 0.43$ | $48.79 \pm 1.86$ | $64.67 \pm 6.75$ | $0.71 \pm 0.07$ |
| | | ODE-NSP | $51.83 \pm 11.61$ | $71.31 \pm 4.10$ | $27.75 \pm 17.41$ | $83.51 \pm 14.07$ | $0.43 \pm 0.12$ |

## State transition Prediction for a Forecasts Horizon of 7 Days on the SocioPatterns and Murcia Datasets

**Table 2:** State transition prediction performance for a forecast horizon of 7 days on the SocioPatterns dataset.

| Model | Epidemiology-informed method | Sensitivity ↑ | Specificity ↑ | Balanced acc. ↑ | AUC ↑ | ECE ↓ |
| --- | --- | --- | --- | --- | --- | --- |
| GAT | None | 65.14 ± 1.33 | 81.54 ± 0.65 | 47.72 ± 1.99 | 85.64 ± 0.62 | 0.09 ± 0.01 |
| | AutoDiff | <u>65.82 ± 2.89</u> | <u>81.95 ± 1.80</u> | <u>48.73 ± 4.33</u> | <u>85.85 ± 4.82</u> | 0.13 ± 0.04 |
| | ODE-NSP | 64.60 ± 1.23 | 81.22 ± 0.63 | 46.90 ± 1.85 | 85.04 ± 0.73 | 0.10 ± 0.03 |
| GraphSAGE | None | **94.50 ± 0.69** | <u>95.17 ± 0.40</u> | **91.76 ± 1.04** | <u>97.84 ± 0.23</u> | <u>0.10 ± 0.01</u> |
| | AutoDiff | 94.15 ± 0.69 | 94.94 ± 0.36 | 91.23 ± 1.03 | 97.77 ± 0.23 | 0.10 ± 0.01 |
| | ODE-NSP | 94.44 ± 0.71 | 95.09 ± 0.40 | 91.65 ± 1.06 | 97.82 ± 0.20 | 0.10 ± 0.01 |
| TGN | None | 83.17 ± 18.72 | 92.80 ± 6.64 | 74.75 ± 28.07 | 93.12 ± 16.46 | 0.09 ± 0.04 |
|  | AutoDiff | <u>94.49 ± 2.03</u> | **95.41 ± 1.26** | <u>91.74 ± 3.05</u> | **98.85 ± 0.55** | **0.07 ± 0.04** |
| | ODE-NSP | 87.54 ± 15.32 | 92.67 ± 5.44 | 81.31 ± 22.98 | 97.97 ± 2.46 | 0.09 ± 0.04 |
| STM-GNN | None | 92.28 ± 7.28 | 94.45 ± 0.61 | 88.42 ± 10.93 | 97.22 ± 1.12 | 0.13 ± 0.02 |
| | AutoDiff | 87.89 ± 17.84 | 92.86 ± 4.95 | 81.83 ± 26.75 | 95.72 ± 2.99 | 0.13 ± 0.02 |
|  | ODE-NSP | <u>93.84 ± 0.93</u> | <u>94.59 ± 0.41</u> | <u>90.77 ± 1.39</u> | <u>97.90 ± 0.32</u> | <u>0.12 ± 0.03</u> |

**Table 3:** State transition prediction performance for a forecast horizons of 7 days on the Murcia dataset.

| Model | Epidemiology-informed method | Sensitivity ↑ | Specificity ↑ | Balanced acc. ↑ | AUC ↑ | ECE ↓ |
| --- | --- | --- | --- | --- | --- | --- |
| GAT | None | $51.70 \pm 11.90$ | $82.14 \pm 3.58$ | $35.61 \pm 15.87$ | $77.62 \pm 6.76$ | $0.12 \pm 0.10$ |
| | AutoDiff | $53.45 \pm 9.34$ | $82.63 \pm 2.85$ | $37.93 \pm 12.46$ | $77.25 \pm 6.16$ | $0.15 \pm 0.18$ |
| | ODE-NSP | $49.74 \pm 7.69$ | $82.63 \pm 2.58$ | $32.99 \pm 10.25$ | $77.31 \pm 6.77$ | $0.12 \pm 0.14$ |
| GraphSAGE | None | $70.62 \pm 3.84$ | $86.15 \pm 1.70$ | $60.82 \pm 5.11$ | $83.70 \pm 2.20$ | $0.06 \pm 0.08$ |
| | AutoDiff | **$71.47 \pm 3.18$** | $86.27 \pm 1.74$ | **$61.97 \pm 4.24$** | $84.15 \pm 2.51$ | $0.06 \pm 0.10$ |
| | ODE-NSP | $69.73 \pm 4.86$ | $85.93 \pm 2.07$ | $59.65 \pm 6.47$ | $82.88 \pm 3.44$ | $0.07 \pm 0.11$ |
| TGN | None | $57.12 \pm 13.89$ | $80.07 \pm 4.87$ | $42.83 \pm 18.53$ | $79.05 \pm 8.67$ | $0.13 \pm 0.09$ |
| | AutoDiff | $57.07 \pm 13.71$ | $80.95 \pm 5.37$ | $42.76 \pm 18.28$ | $77.06 \pm 9.40$ | $0.26 \pm 0.21$ |
| | ODE-NSP | $67.27 \pm 6.07$ | **$87.04 \pm 4.17$** | $56.36 \pm 8.09$ | $83.04 \pm 3.47$ | $0.14 \pm 0.16$ |
| STM-GNN | None | $66.36 \pm 5.16$ | $85.52 \pm 2.31$ | $55.15 \pm 6.88$ | $82.91 \pm 3.42$ | $0.08 \pm 0.09$ |
| | AutoDiff | $63.29 \pm 6.84$ | $84.19 \pm 2.37$ | $51.05 \pm 9.12$ | $79.39 \pm 4.41$ | $0.09 \pm 0.06$ |
| | ODE-NSP | $63.53 \pm 7.29$ | $85.67 \pm 1.42$ | $51.37 \pm 9.72$ | $82.56 \pm 2.90$ | **$0.05 \pm 0.07$** |

## Datasets

### HUG-COVID Dataset

**Table 4:**
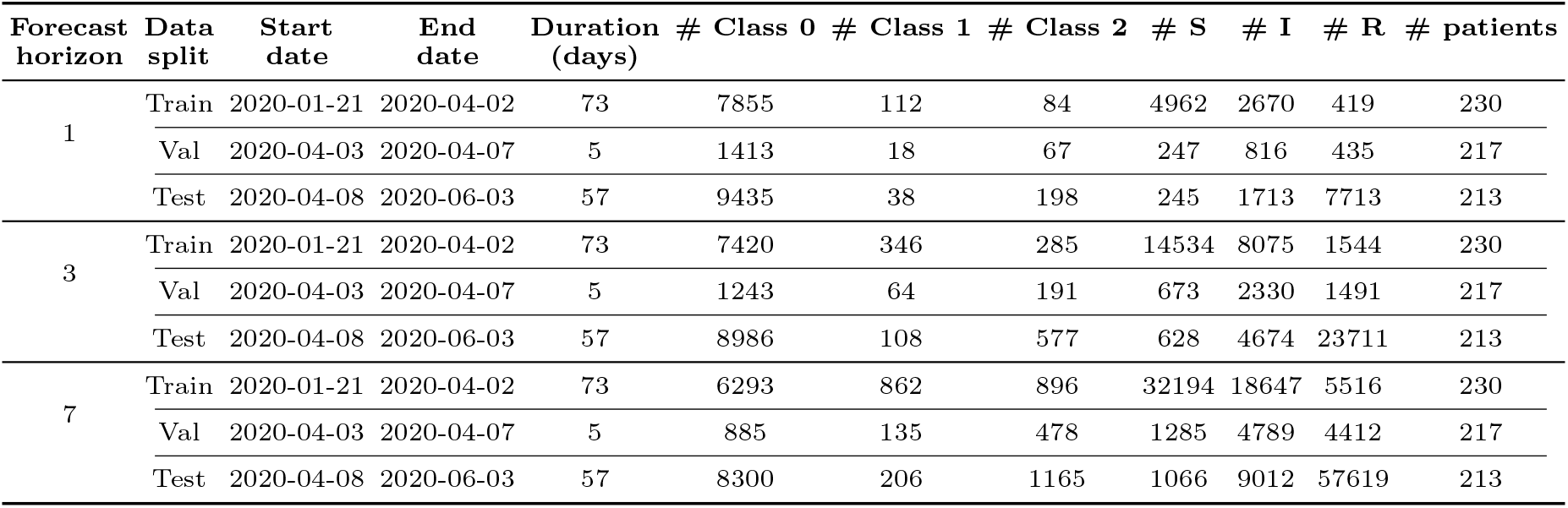
Main HUG-COVID dataset characteristics.

| Forecast horizon | Data split | Start date | End date | Duration (days) | # Class 0 | # Class 1 | # Class 2 | # S | # I | # R | # patients |
| --- | --- | --- | --- | --- | --- | --- | --- | --- | --- | --- | --- |
| 1 | Train | 2020-01-21 | 2020-04-02 | 73 | 7855 | 112 | 84 | 4962 | 2670 | 419 | 230 |
|  | Val | 2020-04-03 | 2020-04-07 | 5 | 1413 | 18 | 67 | 247 | 816 | 435 | 217 |
|  | Test | 2020-04-08 | 2020-06-03 | 57 | 9435 | 38 | 198 | 245 | 1713 | 7713 | 213 |
| 3 | Train | 2020-01-21 | 2020-04-02 | 73 | 7420 | 346 | 285 | 14534 | 8075 | 1544 | 230 |
|  | Val | 2020-04-03 | 2020-04-07 | 5 | 1243 | 64 | 191 | 673 | 2330 | 1491 | 217 |
|  | Test | 2020-04-08 | 2020-06-03 | 57 | 8986 | 108 | 577 | 628 | 4674 | 23711 | 213 |
| 7 | Train | 2020-01-21 | 2020-04-02 | 73 | 6293 | 862 | 896 | 32194 | 18647 | 5516 | 230 |
|  | Val | 2020-04-03 | 2020-04-07 | 5 | 885 | 135 | 478 | 1285 | 4789 | 4412 | 217 |
|  | Test | 2020-04-08 | 2020-06-03 | 57 | 8300 | 206 | 1165 | 1066 | 9012 | 57619 | 213 |

### SocioPatterns Dataset

Given the limited temporal span of the SFHH and Lyon hospital ward datasets, we apply the CONSTR-SH method introduced in [62] to extend each dataset over *N* days. On the resulting dynamic contact networks, we simulate an SEIR infectious disease process using epidemiological parameters consistent with SARS-CoV-2: an incubation period of 5 days, a recovery period of 14 days, and a basic reproduction number of 2.5. As the original datasets provide only interaction data, we simulate patient-level attributes. Static patient features are sampled from binomial distributions (comorbidities: diabetes, stroke, cancer, obesity, smoking), normal distributions (age, subsequently standardized), and Bernoulli distributions (gender and origin, defined as acute care unit, residence, or RSOP).

This procedure yields, for each extended SFHH or Lyon hospital ward dataset, a temporal dataset spanning *N* days in which each individual is associated with (i) dynamic interaction data with other individuals, (ii) a simulated dynamic SEIR state, and (iii) simulated static attributes. From these data, we construct a sequence of bidirectional homogeneous graphs, where nodes represent individuals and an edge is present between two nodes at time *t* if they interacted at that time, edge weights corresponding to the duration of the interaction. Compared with the HUG COVID dataset, these graphs provide more fine-grained information on individual movements and interactions.

Using this framework, we generate 20 independent pseudo-synthetic datasets, each comprising three splits: (i) a training set based on an extended SFHH dataset over 200 days with 100 initially infected individuals, (ii) a validation set based on an extended SFHH dataset over 50 days with 100 initially infected individuals, and (iii) a test set based on the Lyon hospital ward dataset extended over 100 days with 20 initially infected individuals. As for the HUG COVID dataset, we define two prediction tasks—state transitions prediction and epidemiological state prediction—considered for a forecast horizon of *W* = 7 days, resulting in a total of 102,083 samples. Detailed statistics for each split are provided in Tables 5 and 6.

**Table 5:** Main SocioPatterns dataset characteristics (transitions prediction).

| Forecast horizon | Data split | Subdataset | Duration (days) | # Class 0 | # Class 1 | # Class 2 | # patients |
| --- | --- | --- | --- | --- | --- | --- | --- |
| 1 | Train | SFHH | 200 | $79375 \pm 7$ | $419 \pm 7$ | $403 \pm 0$ | 403 |
| | Val | SFHH | 50 | $18927 \pm 16$ | $417 \pm 16$ | $403 \pm 0$ | 403 |
| | Test | Hospital Ward | 100 | $7253 \pm 3$ | $97 \pm 3$ | $75 \pm 0$ | 75 |
| 7 | Train | SFHH | 200 | $72945 \pm 32$ | $2014 \pm 32$ | $2820 \pm 2$ | 403 |
| | Val | SFHH | 50 | $12502 \pm 62$ | $20007 \pm 62$ | $2820 \pm 2$ | 403 |
| | Test | Hospital Ward | 100 | $6026 \pm 12$ | $424 \pm 12$ | $525 \pm 2$ | 75 |

**Table 6:**
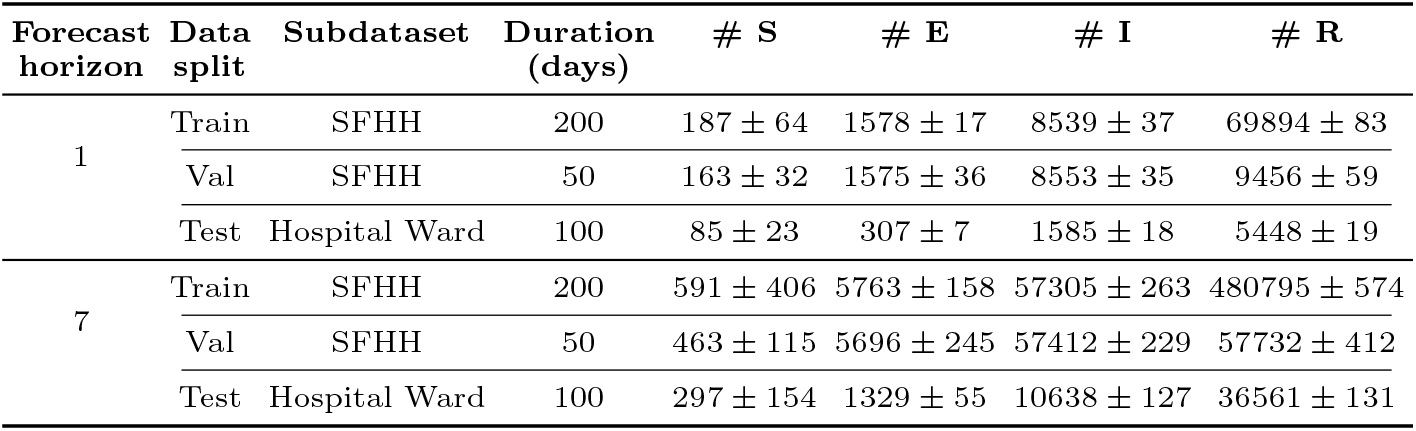
Main SocioPatterns dataset characteristics (states prediction).

| Forecast horizon | Data split | Subdataset | Duration (days) | # S | # E | # I | # R |
| --- | --- | --- | --- | --- | --- | --- | --- |
| 1 | Train | SFHH | 200 | $187 \pm 64$ | $1578 \pm 17$ | $8539 \pm 37$ | $69894 \pm 83$ |
| | Val | SFHH | 50 | $163 \pm 32$ | $1575 \pm 36$ | $8553 \pm 35$ | $9456 \pm 59$ |
| | Test | Hospital Ward | 100 | $85 \pm 23$ | $307 \pm 7$ | $1585 \pm 18$ | $5448 \pm 19$ |
| 7 | Train | SFHH | 200 | $591 \pm 406$ | $5763 \pm 158$ | $57305 \pm 263$ | $480795 \pm 574$ |
| | Val | SFHH | 50 | $463 \pm 115$ | $5696 \pm 245$ | $57412 \pm 229$ | $57732 \pm 412$ |
| | Test | Hospital Ward | 100 | $297 \pm 154$ | $1329 \pm 55$ | $10638 \pm 127$ | $36561 \pm 131$ |

Finally, snapshot timestamps are normalized using a min–max scaler. Edge weights are normalized using Min-Max scaling, followed by logarithmic compression and exponential saturation. All normalization parameters are computed exclusively on the training set to prevent information leakage.

### Murcia Dataset

From each simulated dataset, we extract a temporal graph dataset with a structure closely matching that of the HUG COVID dataset. The main distinction is that place nodes represent not only wards but also surgical and radiology rooms. In addition, this dataset provides a richer set of node-level features. Patient nodes are associated with static features including age, gender, and admission day, as well as dynamic features capturing length of stay (LOS) in the current location, current and previous locations, current service, fine-grained patient location (primarily bed-level), the number of patients co-located with the current patient, and the current SEIRD–NS epidemiological state. Place nodes are described by a static service attribute and dynamic features including contamination status, cleaning status, and the number of patients present. The state transitions prediction and epidemiological state prediction tasks are defined analogously to those used for the HUG COVID and SocioPatterns datasets.

Additionally, each simulated dataset is temporally split into training, validation, and test sets based on fixed time intervals: time steps 0 to 999 (0.91 years) are used for training, time steps 1000 to 1400 (0.37 years) for validation, and time steps 1400 to 2182 (0.71 years) for testing. Detailed statistics for each split are reported in Tables 7 and 8.

**Table 7:**
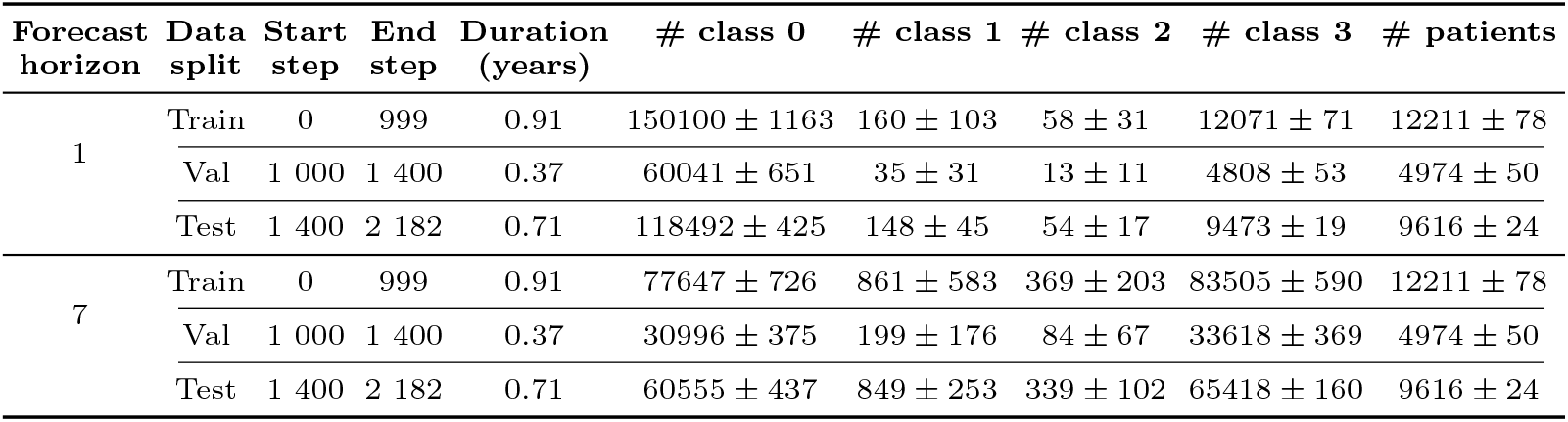
Main Murcia dataset characteristics (transitions prediction).

**Table 8:**
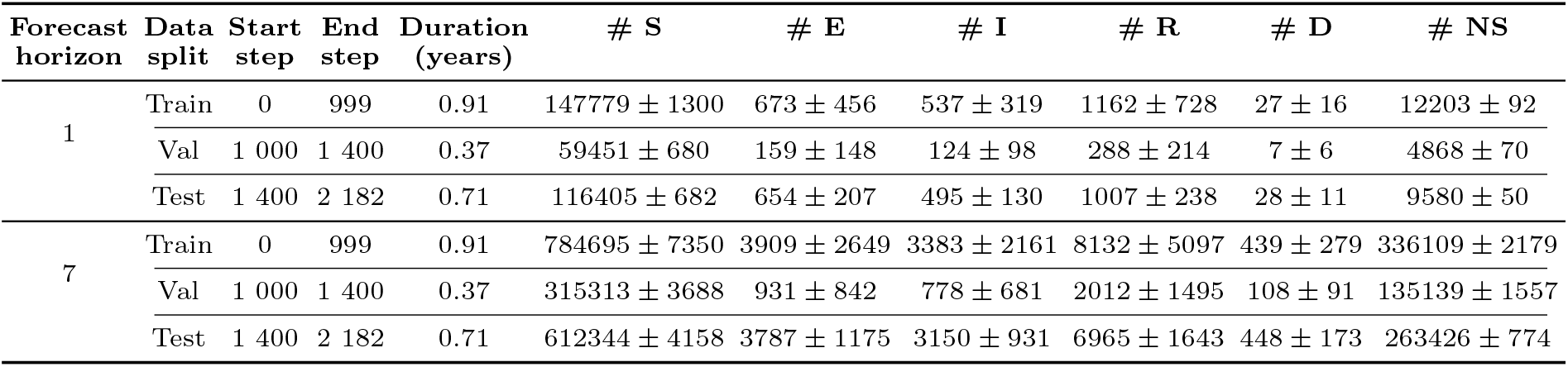
Main Murcia dataset characteristics (states prediction).

Finally, feature normalization is applied using a standard scaler for age and a Min-Max scaler for snapshot timestamps, admission day, LOS, and the number of patients co-located in the same location. All normalization parameters are computed exclusively on the training set to prevent information leakage.

## Internal and external inflows approximations for the SIR model with Immigration/Emigration

The internal and external inflows are approximated using the assumptions in Section 5 and competing risk theory:

- Internal inflows:

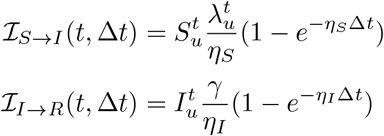
- External inflows:

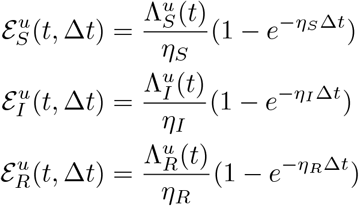

Therefore, the full final state update equations for node *u* are the following:

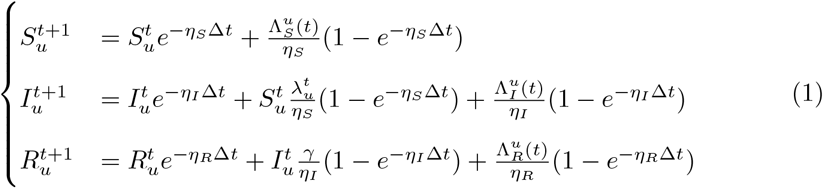

## SEIR and SEIRD-NS Epidemiological Models

### SEIR Model

#### Network-based Model

As for the SIR with immigration/emigration model, we can define the network-based SEIR, for each individual/node *u*,

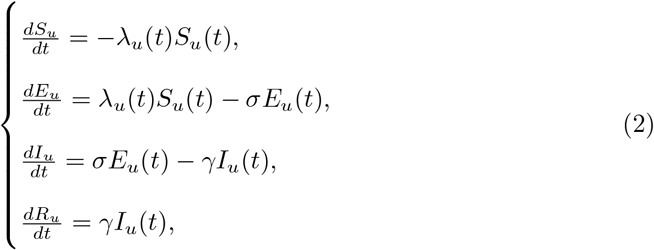

where **S**_*u*_, **E**_*u*_, **I**_*u*_, **R**_*u*_ represent the probability of individual/node *u* being in each compartment at time t, with the constraint **S**_*u*_ + **E**_*u*_ + **I**_*u*_ + **R**_*u*_ = 1, and 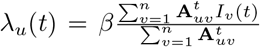 is the local infection hazard (or force of infection), *β* is the infectious/transmission rate, *γ* is the recovery rate (inverse of the average infectious period), and *σ* is the incubation rate (inverse of the average incubation period).

#### Discrete-time hazard/exponential waiting time model

Similarly to the SIR with immigration/emigration model, using the integrating factor method, we can discretize the previous ODEs, under the following assumptions:

- Between times *t* and *t* + Δ*t, λ*_*u*_(*t*) is constant;
- The infection inflow *λ*_*u*_(*t*)*S*_*u*_(*t*) is constant between *t* and *t* + Δ*t*;
- *E*_*u*_ is constant between *t* and *t* + Δ*t*.

Under these assumptions, we have the following discretization of ODEs (2) for the discretization step Δ*t* and individual/node *u*:

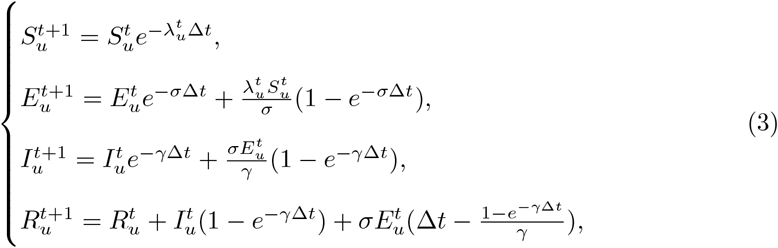

### SEIRD-NS Model

#### Network-based Model

As for the SIR with immigration/emigration and SEIR models, we can define the network-based SEIR, for each individual/node *u*,

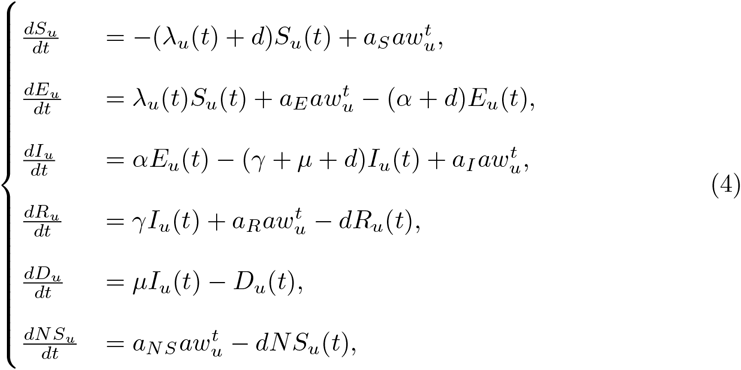

where:

- For node *i* at time *t* let *S*_*u*_, *E*_*u*_, *I*_*u*_, *R*_*u*_, *D*_*u*_, *NS*_*u*_ denote the probabilities (or normalized expected counts) in each compartment;
- *a* is the arrival rate per day;
- *a*_*S*_, *a*_*E*_, *a*_*I*_, *a*_*R*_, and *a*_*NS*_ are the proportions of arrivals in each state;
- *β* is the contact rate or infection rate;
- *µ* is the mortality rate;
- *d* is the discharge rate, defined as the inverse of the length of stay (LOS);
- *α*^−1^ is the average incubation rate;
- *γ* is the recovery rate.
- 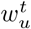 fraction of arrivals landing at node *u* (so ∑_*u*_ *w*_*u*_ = 1). Then arrivals into node *u* in compartment *X* are *a*_*X*_ ∈ {*a*_*S*_, *a*_*E*_, *a*_*I*_, *a*_*R*_, *a*_*NS*_};
- We are going to use 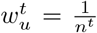 where *n*^*t*^ is the number of nodes at time *t* (for the dynamic case).

#### Discrete-time hazard/exponential waiting time model

Since the arrival parameters *a, a*_*E*_, *a*_*I*_, *a*_*R*_, *a*_*D*_, and *a*_*NS*_ are defined at the population level, they are not included in the individual (node-level) system of ODEs. Under this consideration, and adopting the same assumptions as in the SEIR model, we further assume that 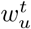 remains constant over the interval [*t, t* + Δ*t*]. The resulting ODE system can then be discretized using the integrating factor method as follows:

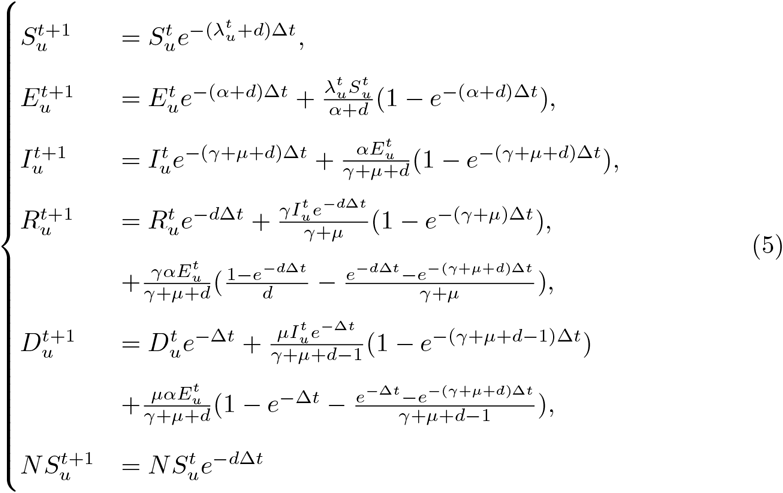

If we consider *I*_*u*_ constant between *t* and *t* + Δ*t*, the equations of *R* and *D* can be simplified as follows:

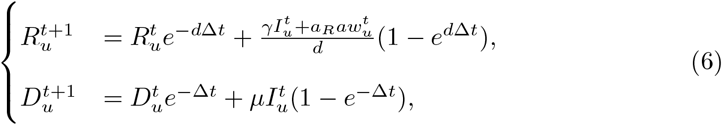

If we ignore the arrivals, we have:

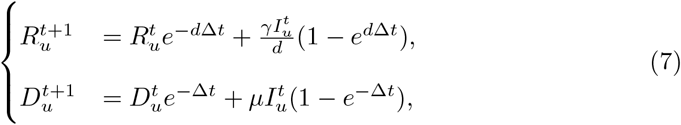

## Temporal Consistency and Transition Constraints

In spatiotemporal epidemiological modeling, the biological progression of a disease imposes strict constraints on state transitions over time. Given the temporal resolution of our hospital dataset (e.g., daily snapshots), certain transitions are physically infeasible within a single time step. To prevent the model from making biologically impossible predictions across the forecast horizon *W*, we introduce a transition constraint loss 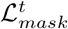.

### Transition Compatibility Matrices

We formalize the valid disease dynamics using a transition compatibility matrix 𝒯 ∈ {0, 1}^*C*×*C*^, where 𝒯_*i,j*_ = 1 if a transition from state *i* at time *t* to state *j* at time *t* + 1 is biologically valid, and 𝒯_*i,j*_ = 0 otherwise.

For the SIR model, with states Susceptible (*S* = 0), Infected (*I* = 1), and Recovered (*R* = 2), the compatibility matrix is defined as:

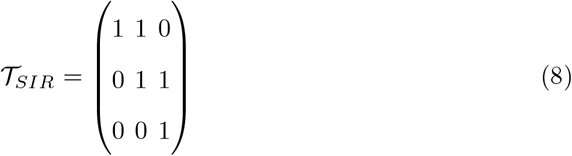

This matrix enforces that a patient cannot skip the infection stage (*S* → *R*) and cannot revert to a previous state (*I* → *S* or *R* → *I*).

For the SEIR model, which includes an Exposed (*E* = 1) state, the matrix is:

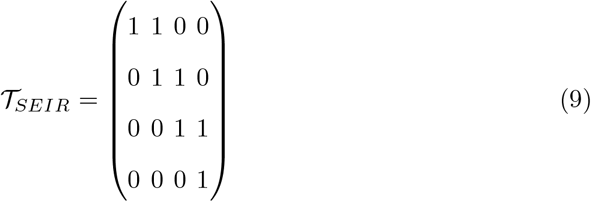

For the SEIRD-NS model, which includes an Exposed (*E* = 1) state, the matrix is:

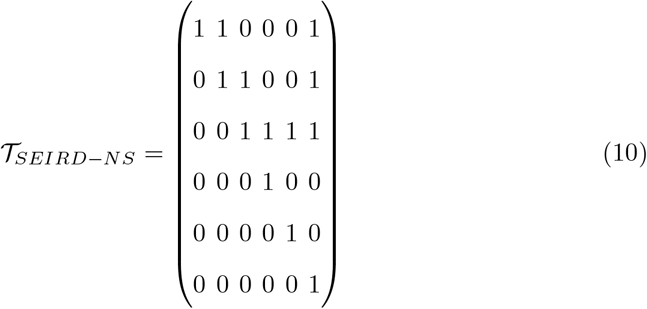

## Epidemic models weights initialization

For the **HUG-COVD** dataset, we used the same initialization for both models ℱ_*ODE*_ (AutoDiff) and *p*_*θ*_ (ODE-NSP): *β* = 0.5, *γ* = 0.1, Λ_*S*_ = 0.02, Λ_*I*_ = 0.001, Λ_*R*_ = 0.001, *µ*_*S*_ = 0.01, *µ*_*I*_ = 0.02, *µ*_*R*_ = 0.005, and *ϵ* = 0.001.

For the **SocioPatterns** dataset, we also used the same initialization for both models ℱ_*ODE*_ (AutoDiff) and *p*_*θ*_ (ODE-NSP): *β* = 0.5, *σ* = 1*/*7, and *γ* = 1*/*12.

For the **Murcia** dataset, we used the different initialization for ℱ_*ODE*_ (AutoDiff) and *p*_*θ*_ (ODE-NSP). Indeed, ℱ_*ODE*_ (AutoDiff) requires additional parameters thatare not used for *p*_*θ*_ (ODE-NSP). Thus, for ℱ_*ODE*_ and *p*_*θ*_ we used: *β* = 1*/*0.675, *d* = 1*/*4.254, *α* = 1*/*2.5, *µ* = 0.027, and *γ* = 1 − *µ*. In addition, for ℱ_*ODE*_ we used: *a* = 18.603, *a*_*S*_ = 0.997, *a*_*E*_ = 1*e*^−7^, *a*_*I*_ = 0.002, *a*_*R*_ = 1*e*^−7^, and *a*_*NS*_ = 0.001.

## References

[1] Tran Quoc, V., Nguyen Thi Ngoc, D., Nguyen Hoang, T., Vu Thi, H., Tong Duc, M., Do Pham Nguyet, T., Nguyen Van, T., Ho Ngoc, D., Vu Son, G., Bui Duc, T.: Predicting Antibiotic Resistance in ICUs Patients by Applying Machine Learning in Vietnam. Infection and Drug Resistance 16, 5535–5546 (2023) 10.2147/IDR.S415885 . Accessed 2025-09-04

[2] Wang, J., Wang, G., Wang, Y., Wang, Y.: Development and evaluation of a model for predicting the risk of healthcare-associated infections in patients admitted to intensive care units. Frontiers in Public Health 12, 1444176 (2024) 10.3389/fpubh.2024.1444176 . Accessed 2025-09-04

[3] Murri, R., De Angelis, G., Antenucci, L., Fiori, B., Rinaldi, R., Fantoni, M., Damiani, A., Patarnello, S., Sanguinetti, M., Valentini, V., Posteraro, B., Masciocchi, C.: A Machine Learning Predictive Model of Bloodstream Infection in Hospitalized Patients. Diagnostics 14(4), 445 (2024) 10.3390/diagnostics14040445 . Accessed 2025-09-04

[4] Liu, J.-Y., Dickter, J.K.: Nosocomial Infections: A History of Hospital-Acquired Infections. Gastrointestinal Endoscopy Clinics of North America 30(4), 637–652 (2020) 10.1016/j.giec.2020.06.001

[5] Longa, A., Lachi, V., Santin, G., Bianchini, M., Lepri, B., Lio, P., scarselli Passerini, A.: Graph neural networks for temporal graphs: State of the art, open challenges, and opportunities. Transactions on Machine Learning Research (2023)

[6] Zheng, Y., Yi, L., Wei, Z.: A survey of dynamic graph neural networks. Frontiers of Computer Science 19(6), 196323 (2024) 10.1007/s11704-024-3853-2 . Accessed 2025-12-15

[7] Gouareb, R., Bornet, A., Proios, D., Pereira, S.G., Teodoro, D.: Detection of patients at risk of multidrug-resistant enterobacteriaceae infection using graph neural networks: A retrospective study. Health Data Science 3, 0099 (2023) 10.34133/hds.0099 https://spj.science.org/doi/pdf/10.34133/hds.0099

[8] Geissbuhler, D., Bornet, A., Marques, C., Anjos, A., Pereira, S., Teodoro, D.: Stm-gnn: Space-time-and-memory graph neural networks for predicting multi-drug resistance risks in dynamic patient networks. In: Bellazzi, R., Juarez Herrero, J.M., Sacchi, L., Zupan, B. (eds.) Artificial Intelligence in Medicine, pp. 160–169. Springer, Cham (2025)

[9] Pareja, A., Domeniconi, G., Chen, J., Ma, T., Suzumura, T., Kanezashi, H., Kaler, T., Schardl, T.B., Leiserson, C.E.: EvolveGCN: Evolving graph convolutional networks for dynamic graphs. In: Proceedings of the Thirty-Fourth AAAI Conference on Artificial Intelligence (2020)

[10] Sankar, A., Wu, Y., Gou, L., Zhang, W., Yang, H.: Dysat: Deep neural representation learning on dynamic graphs via self-attention networks. In: Proceedings of the 13th International Conference on Web Search and Data Mining. WSDM ‘20, pp. 519–527. Association for Computing Machinery, New York, NY, USA (2020). https://doi.org.10.1145/3336191.3371845. 10.1145/3336191.3371845

[11] Yu, B., Yin, H., Zhu, Z.: Spatio-temporal graph convolutional networks: A deep learning framework for traffic forecasting. In: Proceedings of the Twenty-Seventh International Joint Conference on Artificial Intelligence, IJCAI-18, pp. 3634–3640. International Joint Conferences on Artificial Intelligence Organization, ??? (2018). 10.24963/ijcai.2018/505. https://doi.org/10.24963/ijcai.2018/505

[12] Zhao, L., Song, Y., Zhang, C., Liu, Y., Wang, P., Lin, T., Deng, M., Li, H.: T-gcn: A temporal graph convolutional network for traffic prediction. IEEE Transactions on Intelligent Transportation Systems 21(9), 3848–3858 (2020) 10.1109/TITS.2019.2935152

[13] Wu, Z., Pan, S., Long, G., Jiang, J., Zhang, C.: Graph wavenet for deep spatial-temporal graph modeling. In: Proceedings of the 28th International Joint Conference on Artificial Intelligence. IJCAI’19, pp. 1907–1913. AAAI Press, ??? (2019)

[14] Bai, L., Yao, L., Li, C., Wang, X., Wang, C.: Adaptive graph convolutional recurrent network for traffic forecasting. In: Proceedings of the 34th International Conference on Neural Information Processing Systems. NIPS ‘20. Curran Associates Inc., Red Hook, NY, USA (2020)

[15] Qin, X., Sheikh, N., Lei, C., Reinwald, B., Domeniconi, G.: Seign: A simple and efficient graph neural network for large dynamic graphs. In: 2023 IEEE 39th International Conference on Data Engineering (ICDE), pp. 2850–2863 (2023). 10.1109/ICDE55515.2023.00218

[16] Han, M.: Learning dynamic graphs via tensorized and lightweight graph convolutional networks (2025) arXiv:2504.15613 [cs.LG]

[17] Trivedi, R., Farajtabar, M., Biswal, P., Zha, H.: Dyrep: Learning representations over dynamic graphs. In: International Conference on Learning Representations (2019). https://openreview.net/forum?id=HyePrhR5KX

[18] Rossi, E., Chamberlain, B., Frasca, F., Eynard, D., Monti, F., Bronstein, M.: Temporal graph networks for deep learning on dynamic graphs. In: ICML 2020 Workshop on Graph Representation Learning (2020)

[19] Qarkaxhija, L., Perri, V., Scholtes, I.: De bruijn goes neural: Causality-aware graph neural networks for time series data on dynamic graphs. In: Rieck, B., Pascanu, R. (eds.) Proceedings of the First Learning on Graphs Conference. Proceedings of Machine Learning Research, vol. 198, pp. 51–15121. PMLR, ??? (2022). https://proceedings.mlr.press/v198/qarkaxhija22a.html

[20] Biparva, M., Karimi, R., Faez, F., Zhang, Y.: Todyformer: Towards holistic dynamic graph transformers with structure-aware tokenization (2024)

[21] Peng, J., Wei, Z., Ye, Y.: Tidformer: Exploiting temporal and interactive dynamics makes a great dynamic graph transformer. In: Proceedings of the 31st ACM SIGKDD Conference on Knowledge Discovery and Data Mining V.2. KDD ‘25, pp. 2245–2256. Association for Computing Machinery, New York, NY, USA (2025). 10.1145/3711896.3737155. https://doi.org/10.1145/3711896.3737155

[22] Zhu, X., Zhang, Y., Ying, H., Chi, H., Sun, G., Zeng, L.: Modeling epidemic dynamics using Graph Attention based Spatial Temporal networks. PLOS ONE 19(7), 0307159 (2024) 10.1371/journal.pone.0307159 . Publisher: Public Library of Science. Accessed 2025-12-15

[23] Goyal, P., Kamra, N., He, X., Liu, Y.: Dyngem: Deep embedding method for dynamic graphs. arXiv preprint arXiv:1805.11273 (2018)

[24] Zhou, L., Yang, Y., Ren, X., Wu, F., Zhuang, Y.: Dynamic network embedding by modeling triadic closure process. In: Proceedings of the Thirty-Second AAAI Conference on Artificial Intelligence and Thirtieth Innovative Applications of Artificial Intelligence Conference and Eighth AAAI Symposium on Educational Advances in Artificial Intelligence. AAAI’18/IAAI’18/EAAI’18. AAAI Press, ??? (2018)

[25] Kazemi, S.M., Goel, R., Jain, K., Kobyzev, I., Sethi, A., Forsyth, P., Poupart, P.: Representation learning for dynamic graphs: a survey. J. Mach. Learn. Res. 21(1) (2020)

[26] Skarding, J., Gabrys, B., Musial, K.: Foundations and modeling of dynamic networks using dynamic graph neural networks: A survey. IEEE Access 9, 79143–79168 (2021) 10.1109/ACCESS.2021.3082932

[27] Huang, S., Poursafaei, F., Danovitch, J., Fey, M., Hu, W., Rossi, E., Leskovec, J., Bronstein, M.M., Rabusseau, G., Rabbany, R.: Temporal graph benchmark for machine learning on temporal graphs. In: Thirty-seventh Conference on Neural Information Processing Systems Datasets and Benchmarks Track (2023). https://openreview.net/forum?id=qG7IkQ7IBO

[28] Hu, N., Zhang, D., Xie, K., Liang, W., Hsieh, M.-Y.: Graph learning-based spatial-temporal graph convolutional neural networks for traffic forecasting. Connection Science 34(1), 429–448 (2022) 10.1080/09540091.2021.2006607

[29] Zhang, A.: Dynamic graph convolutional networks with Temporal representation learning for traffic flow prediction. Scientific Reports 15(1), 17270 (2025) 10.1038/s41598-025-01696-7 . Accessed 2025-12-15

[30] Wang, L., Han, M.: Neighborhood overlap-aware high-order graph neural network for dynamic graph learning. In: 9th International Conference on Electronic Information Technology and Computer Engineering (EITCE 2025), vol. 2025, pp. 101–105 (2025). 10.1049/icp.2025.2855

[31] Raissi, M., Perdikaris, P., Karniadakis, G.E.: Physics-informed neural networks: A deep learning framework for solving forward and inverse problems involving nonlinear partial differential equations. Journal of Computational Physics 378, 686–707 (2019) 10.1016/j.jcp.2018.10.045

[32] Shaier, S., Raissi, M., Seshaiyer, P.: Data-Driven Approaches for Predicting Spread of Infectious Diseases Through DINNs: Disease Informed Neural Networks. Letters in Biomathematics 9(1), 71–105 (2022) 10.30707/LiB9.1.1681913305.249476 . Accessed 2025-09-19

[33] Tronstad, M.: A Physics-Informed Deep Learning Framework for Solving Inverse Problems in Epidemiology. Masther Thesis, KTH Royal Institute of Technology (2022). https://urn.kb.se/resolve?urn=urn:nbn:se:kth:diva-321233 Accessed 2025-09-19

[34] Rodríguez, A., Cui, J., Ramakrishnan, N., Adhikari, B., Prakash, B.A.: EINNs: Epidemiologically-Informed Neural Networks. Proceedings of the AAAI Conference on Artificial Intelligence 37(12), 14453–14460 (2023) 10.1609/aaai.v37i12.26690 . Accessed 2025-09-19

[35] Millevoi, C., Pasetto, D., Ferronato, M.: A Physics-Informed Neural Network approach for compartmental epidemiological models. PLOS Computational Biology 20(9), 1012387 (2024) 10.1371/journal.pcbi.1012387 . Accessed 2025-09-19

[36] Cheng, C., Aruchunan, E., Noor Aziz, M.H.: Leveraging dynamics informed neural networks for predictive modeling of COVID-19 spread: a hybrid SEIRV-DNNs approach. Scientific Reports 15(1), 2043 (2025) 10.1038/s41598-025-85440-1 . Accessed 2025-09-19

[37] Naser, M.Z.: Fundamental flaws of physics-informed neural networks and explain-ability methods in engineering systems. Computers and Industrial Engineering 212, 111704 (2026) 10.1016/j.cie.2025.111704

[38] Satyadharma, A., Chern, M.-J., Kan, H.-C., Harinaldi Julian, J.: Assessing physics-informed neural network performance with sparse noisy velocity data. Physics of Fluids 36(10), 103619 (2024) 10.1063/5.0213522

[39] Navarin, N., Frazzetto, P., Pasa, L., Verzelli, P., Visentin, F., Sperduti, A., Alippi, C.: Physics-Informed Graph Neural Cellular Automata: an Application to Compartmental Modelling. In: 2024 International Joint Conference on Neural Networks (IJCNN), pp. 1–9 (2024). 10.1109/IJCNN60899.2024.10650578 . ISSN: 2161-4407

[40] Han, S., Stelz, L., Sokolowski, T.R., Zhou, K., Stöcker, H.: Unifying Physics- and Data-Driven Modeling via Novel Causal Spatiotemporal Graph Neural Network for Interpretable Epidemic Forecasting. arXiv. arXiv:2504.05140 (2025). 10.48550/arXiv.2504.05140. http://arxiv.org/abs/2504.05140 Accessed 2025-09-19

[41] Diao, H., Wen, G., Xu, S., Hu, W., Nian, F.: Physics-Informed GNN Epidemic Forecasting via Double-Layer Dynamic Model. IEEE Transactions on Computational Social Systems, 1–14 (2025) 10.1109/TCSS.2025.3555723 . Accessed 2025-09-19

[42] Jeong, B., Lee, Y.J., Han, C.E.: A simple yet effective approach for predicting disease spread using mathematically-inspired diffusion-informed neural networks. Scientific Reports 15(1), 15000 (2025) 10.1038/s41598-025-98398-x . Accessed 2025-09-19

[43] Zheng, Y., Jiang, W., Zhou, A., Hung, N.Q.V., Zhan, C., Chen, T.: Epidemiology-informed Graph Neural Network for Heterogeneity-aware Epidemic Forecasting. arXiv. arXiv:2411.17372 (2024). 10.48550/arXiv.2411.17372 . http://arxiv.org/abs/2411.17372 Accessed 2025-09-19

[44] Mallick, T., Balaprakash, P., Macfarlane, J.: Deep-Ensemble-Based Uncertainty Quantification in Spatiotemporal Graph Neural Networks for Traffic Forecasting (2022). https://arxiv.org/abs/2204.01618

[45] La Gatta, V., Moscato, V., Postiglione, M., Sperlí, G.: An Epidemiological Neural Network Exploiting Dynamic Graph Structured Data Applied to the COVID-19 Outbreak. IEEE Transactions on Big Data 7(1), 45–55 (2021) 10.1109/TBDATA.2020.3032755 . Accessed 2025-09-19

[46] Liu, M., Liu, Y., Liu, J.: Epidemiology-aware Deep Learning for Infectious Disease Dynamics Prediction. In: Proceedings of the 32nd ACM International Conference on Information and Knowledge Management, pp. 4084–4088. ACM, Birmingham United Kingdom (2023). 10.1145/3583780.3615139 . https://dl.acm.org/doi/10.1145/3583780.3615139 Accessed 2025-09-19

[47] Sun, C., Fang, R., Salemi, M., Prosperi, M., Magalis, B.R.: DeepDynaForecast: Phylogenetic-informed graph deep learning for epidemic transmission dynamic prediction. PLOS Computational Biology 20(4), 1011351 (2024) 10.1371/journal.pcbi.1011351 . Accessed 2025-09-19

[48] Alfas, M., Kumar, M., Shriyam, S., Kumar, S.: An Efficient Framework for Epidemiological Parameter Estimation via Graph Reduction and Graph Neural Networks. ACM Transactions on Knowledge Discovery from Data 19(6), 1–29 (2025) 10.1145/3736727 . Accessed 2025-09-19

[49] Tan, C.W., Yu, P.-D., Chen, S., Poor, H.V.: DeepTrace: Learning to Optimize Contact Tracing in Epidemic Networks With Graph Neural Networks. IEEE Transactions on Signal and Information Processing over Networks 11, 97–113 (2025) 10.1109/TSIPN.2025.3530346 . Accessed 2025-09-19

[50] Wan, G., Liu, Z., Shan, X., Lau, M.S., Prakash, B.A., Jin, W.: EARTH: Epidemiology-aware neural ODE with continuous disease transmission graph. In: Forty-second International Conference on Machine Learning (2025). https://openreview.net/forum?id=Cnfogmxymj

[51] Fritz, C., Dorigatti, E., Rügamer, D.: Combining graph neural networks and spatio-temporal disease models to improve the prediction of weekly COVID-19 cases in Germany. Scientific Reports 12(1), 3930 (2022) 10.1038/s41598-022-07757-5 . Accessed 2025-09-18

[52] Park, J.: Graph-based epidemic model using spatio-temporal data. Master’s thesis, Seoul National University, Seoul, South Korea (June 2025). https://s-space.snu.ac.kr/handle/10371/215342

[53] Myall, A., Price, J.R., Peach, R.L., Abbas, M., Mookerjee, S., Zhu, N., Ahmad, I., Ming, D., Ramzan, F., Teixeira, D., Graf, C., Weiße, A.Y., Harbarth, S., Holmes, A., Barahona, M.: Prediction of hospital-onset COVID-19 infections using dynamic networks of patient contact: an international retrospective cohort study. The Lancet. Digital Health 4(8), 573–583 (2022) 10.1016/S2589-7500(22)00093-0

[54] G’enois, M., Barrat, A.: Can co-location be used as a proxy for face-to-face contacts? EPJ Data Science 7(1), 11 (2018) 10.1140/epjds/s13688-018-0140-1

[55] Vanhems, P., Barrat, A., Cattuto, C., Pinton, J.-F., Khanafer, N., Regis, C., Kim, B.-a., Comte, B., Voirin, N.: Estimating potential infection transmission routes in hospital wards using wearable proximity sensors. PLoS ONE 8(9), 73970 (2013) 10.1371/journal.pone.0073970

[56] Kim, D., Canovas-Segura, B., Jimeno-Almazán, A., Campos, M., Juarez, J.M.: Spatial-temporal simulation for hospital infection spread and outbreaks of Clostridioides difficile. Scientific Reports 13(1), 20022 (2023) 10.1038/s41598-023-47296-1 . Accessed 2026-01-05

[57] Liu, Y., Rocklöv, J.: The reproductive number of the Delta variant of SARS-CoV-2 is far higher compared to the ancestral SARS-CoV-2 virus. Journal of Travel Medicine 28(7), 124 (2021) 10.1093/jtm/taab124 . Accessed 2026-03-12

[58] Liu, T., Huang, J., He, Z., Zhang, Y., Yan, N., Zhang, C.J.P., Ming, W.-K.: A real-world data validation of the value of early-stage SIR modelling to public health. Scientific Reports 13(1), 9164 (2023) 10.1038/s41598-023-36386-9 . Accessed 2026-03-12

[59] Veličković, P., Cucurull, G., Casanova, A., Romero, A., Liò, P., Bengio, Y.: Graph attention networks. In: International Conference on Learning Representations (2018)

[60] Hamilton, W.L., Ying, R., Leskovec, J.: Inductive representation learning on large graphs. In: Proceedings of the 31st International Conference on Neural Information Processing Systems. NIPS’17, pp. 1025–1035. Curran Associates Inc., Red Hook, NY, USA (2017)

[61] Akiba, T., Sano, S., Yanase, T., Ohta, T., Koyama, M.: Optuna: A next-generation hyperparameter optimization framework. In: Proceedings of the 25th ACM SIGKDD International Conference on Knowledge Discovery and Data Mining (2019)

[62] Stehlé, J., Voirin, N., Barrat, A., Cattuto, C., Colizza, V., Isella, L., Regis, C., Pinton, J., Khanafer, N., Van den Broeck, W., Vanhems, P.: Simulation of an seir infectious disease model on the dynamic contact network of conference attendees. BMC Medicine 9(87) (2011) 10.1186/1741-7015-9-87

